# MVP-VSASL: measuring MicroVascular Pulsatility using Velocity-Selective Arterial Spin Labeling

**DOI:** 10.1101/2024.06.21.24309261

**Authors:** Conan Chen, Ryan A. Barnes, Katherine J. Bangen, Fei Han, Josef Pfeuffer, Eric C. Wong, Thomas T. Liu, Divya S. Bolar

**Author notes:** **Correspondence To:** Conan Chen, Center for Functional MRI, 9500 Gilman Dr, MC 0677, La Jolla, CA 92093.

## Abstract

1

**Purpose:** By leveraging the small-vessel specificity of velocity-selective arterial spin labeling (VSASL), we present a novel technique for measuring cerebral MicroVascular Pulsatility named MVP-VSASL.

**Theory and Methods:** We present a theoretical model relating the pulsatile, cerebral blood flow-driven VSASL signal to the microvascular pulsatility index (PI), a widely used metric for quantifying cardiac-dependent fluctuations. The model describes the dependence of PI on bolus duration *τ* (an adjustable VSASL sequence parameter) and provides guidance for selecting a value of *τ* that maximizes the SNR of the PI measurement. The model predictions were assessed in humans using data acquired with retrospectively cardiac-gated VSASL sequences over a broad range of *τ* values. In vivo measurements were also used to demonstrate the feasibility of whole-brain voxel-wise PI mapping, assess intrasession repeatability of the PI measurement, and illustrate the potential of this method to explore an association with age.

**Results:** The theoretical model showed excellent agreement to the empirical data in a gray matter region of interest (average R^2^ value of 0.898 *±* 0.107 across six subjects). We further showed excellent intrasession repeatability of the pulsatility measurement (ICC = 0.960, *p<* 0.001) and the potential to characterize associations with age (*r* = 0.554, *p* = 0.021).

**Conclusion:** We have introduced a novel, VSASL-based cerebral microvascular pulsatility technique, which may facilitate investigation of cognitive disorders where damage to the microvasculature has been implicated.

## 2 Introduction

Pulsatile blood flow driven by the cardiac cycle has been increasingly linked to structural damage in the cerebral microvasculature^1–4^ and cognitive disorders such as mild cognitive impairment, Alzheimer’s disease, and other dementias^1–7^. In young, healthy subjects, this flow pulsatility is dampened by compliant arteries as the pulse wave travels toward the distal microvasculature and brain parenchyma. However, if this dampening is insufficient, for example, due to pathologic changes in the vasculature, then excess pulsatile energy will reach the smaller vessels, where the resulting exposure and subsequent damage is thought to be a precursor to the aforementioned cognitive disorders^3,8^. There has been much work assessing the pulsatility^4,8–12^ and vessel wall compliance^10,13–17^ at the large cerebral arteries. However, techniques measuring pulsatility in the microvasculature itself, the site where damage is thought to primarily occur, have received less attention. Such techniques will be crucial to clarifying the exact mechanistic links between microvascular pulsatility, tissue damage, and eventual neurodegeneration, which are not yet fully understood. Furthermore, since neurovascular factors such as flow pulsatility are viewed as early and modifiable risk factors contributing to these disorders, the ability to measure these biomarkers may be important for developing strategies for early detection and intervention^5–7,18^.

Historically, microvascular pulsatility has been challenging to measure due to the very small size of microvascular vessels and their relatively slow flow. Recent approaches using phase contrast MRI have leveraged an ultra-high 7T field strength to measure pulsatility in small perforating arteries^19–22^. Additional advances to data acquisition (higher temporal resolution^20^, dual velocity encoding^22^) and post-processing (automated vessel detection^20^) have improved the usability and robustness of the technique even further. However, this approach remains challenging at the lower field strengths common on clinical scanners, which limits its potential for clinical translation. As an alternative approach, we utilize a technique called velocity-selective arterial spin labeling (VSASL)^23^ to measure microvascular pulsatility. VSASL can be performed on clinical 3T scanners with a simple whole-brain scan prescription and has the potential to generate cerebral microvascular pulsatility maps on a voxel-wise basis.

VSASL is a variant of arterial spin labeling (ASL), a family of methods used to measure perfusion by magnetically labeling a bolus of arterial blood, allowing the bolus to flow into the microvasculature or tissue of interest, and then acquiring images sensitized to the blood flow^24^. While the traditional brain ASL variants generate the magnetic label within the feeding extracranial carotid and vertebral arteries, VSASL is unique in its ability to generate the label in smaller, more distal arteries^23^. In VSASL, the user-specified sequence parameter of cutoff velocity (*v*_cut_) determines the location along the vasculature where the leading and trailing edges of the bolus are defined, as illustrated in Figure 1. Using a typical setting of *v*_cut_ = 2 cm/s^23^, the sequence will define the boundary of the labeled bolus in small arterioles with vessel diameters of about 50 µm^25^, which lies within the range of vessel diameters in the microvasculature. In the standard VSASL sequence, a velocity-selective (VS) label/control module (LCM) will first label blood flowing faster than *v*_cut_, thus defining the leading edge of the labeled bolus (Figure 1A). This is followed by a delay corresponding to the bolus duration *τ* (also a user-specified sequence parameter)^23^, during which the bolus flows distally toward its target tissue while decelerating below *v*_cut_ in the process (Figure 1B). A second VS module called the vascular crushing module (VCM) is then applied to saturate remaining labeled blood still flowing faster than *v*_cut_, thus defining the trailing edge of the labeled bolus (Figure 1C). The labeled bolus signal is proportional to the volume of blood that flows across the *v*_cut_ boundary in the time between LCM and VCM, thus making the labeled bolus signal (i.e. VSASL signal) sensitive to flow rate and its variation across the cardiac cycle.

**Figure 1:**
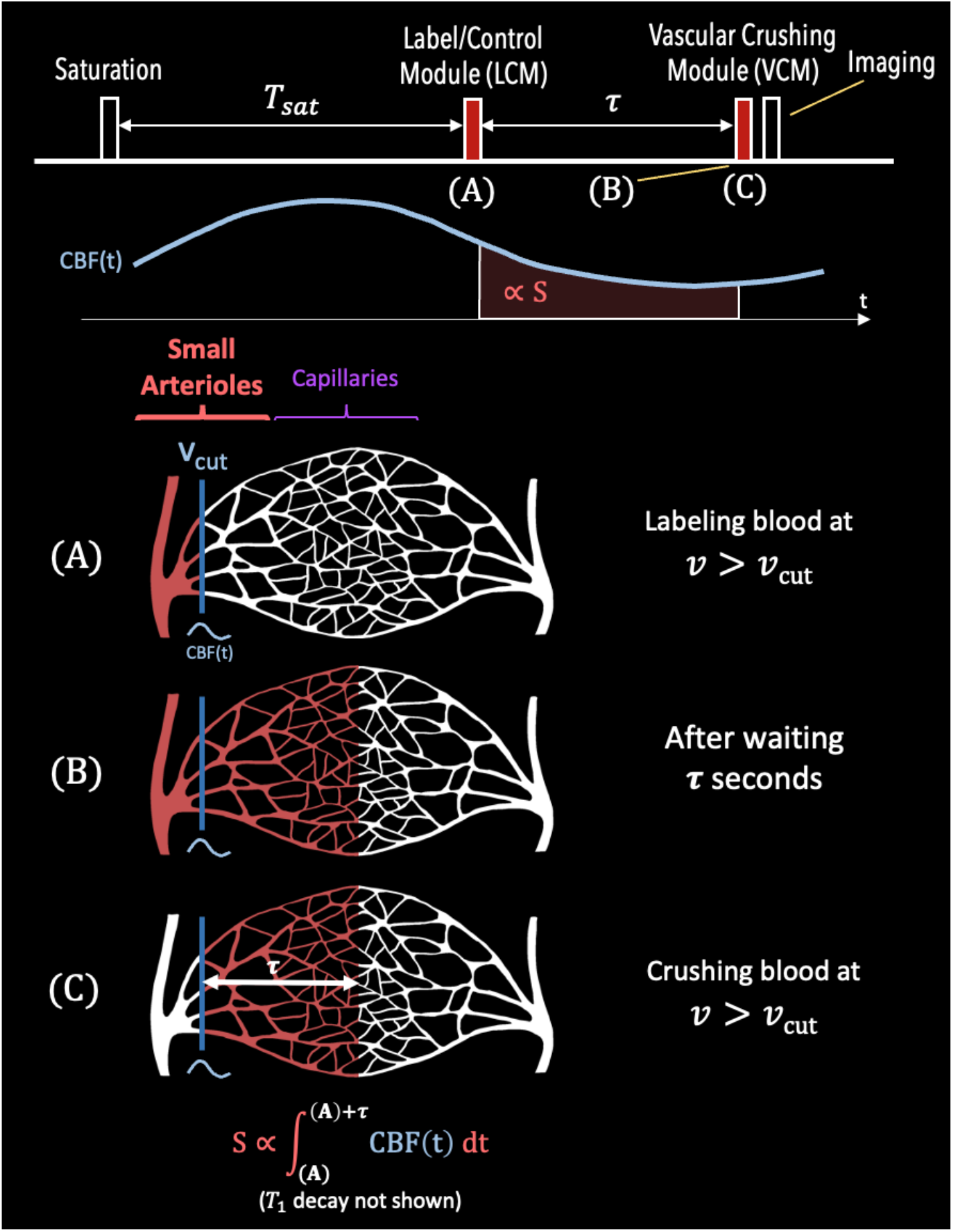
Illustration of the VSASL labeling process. The top most diagram shows the VSASL pulse sequence, with events (A)-(C) corresponding to the sub-figures below. Below the sequence diagram is an example CBF(*t*) waveform, with the area shaded in red representing the VSASL signal *S*. Events (A)-(C) are described as follows: (A) Blood signal flowing faster than the cutoff velocity *v*_cut_ is labeled by the LCM, which defines the leading edge of the bolus. Since *v*_cut_ is specified to the velocity in small arterioles (using a typical *v*_cut_ = 2 cm/s for our study), this leading edge is defined in the microvascular regime. (B) After labeling, there is a waiting period of *τ* seconds, where *τ* is the user-specified bolus duration. During this period, blood continues to flow downstream, past the *v*_cut_ boundary and toward the capillaries and tissue. (C) Blood signal flowing faster than *v*_cut_ is crushed by the VCM. This action defines the trailing edge of the bolus, and thus fully defines the temporal duration of the bolus as *τ*, with the image then acquired shortly thereafter.

Previous work by Franklin et al. using a single VS labeling module alone (the LCM) measured fluctuations of up to 36% in the amount of arterial label generated^26^. This measurement was made in an arterial ROI and was weighted by macrovascular blood volume and flow, since the LCM was applied without an accompanying VCM. Nevertheless, the results suggested the potential of VSASL to measure effects of cardiac pulsations. In this study, we use a standard VSASL sequence design that includes both the LCM and VCM^23^ to specifically achieve microvascular blood flow weighting. By then using retrospective cardiac gating and leveraging the microvascular specificity of VSASL, we realize the potential of measuring blood flow pulsatility in the microvasculature. We dub this technique MicroVascular Pulsatility using VSASL (MVP-VSASL).

We first present theory relating the pulsatile VSASL signal to the underlying pulsatile blood flow, and then examine the pulsatility of the VSASL signal using the pulsatility index (PI) metric. We derive a theoretical model describing the dependence of PI on bolus duration *τ*, and use the model to determine a theoretically optimal *τ* value for maximizing the SNR of the PI measurement. Using experimental VSASL data acquired in human subjects with a broad range of ages and heart rates, we then validate the predicted *τ*-dependence of PI and assess the intrasession test-retest repeatability of the PI measurement. In addition, we examine the association between pulsatility and age and demonstrate the feasibility of a voxel-wise pulsatility mapping approach that may facilitate regional pulsatility measurements in future applications.

## 3 Theory

### 3.1 Pulsatility of VSASL Signal

The control-label subtraction signal (denoted *S*) from a VSASL scan represents the signal of the labeled blood being delivered to the microvasculature. This signal *S* is typically modeled as being proportional to CBF_0_ · *τ* · exp (−*τ*/*T*_1*b*_), where CBF_0_ reflects uniform (non-pulsatile) cerebral blood flow, *τ* is bolus duration, and exp (− *τ/T*_1*b*_) is the *T*_1_ decay weighting factor (where *T*_1*b*_ is the *T*_1_ of blood)^23^.

In the case of time-varying, pulsatile blood flow, the product CBF_0_ · *τ* becomes an integration of CBF(*t*) over the duration of the bolus, and the continuous-time VSASL signal *S*(*t*) can be described as:

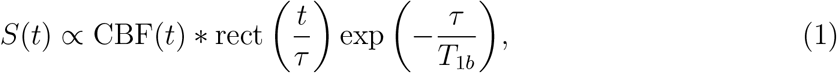

where *\** denotes convolution and rect(*t/τ*) is a rectangular function of width *τ* representing the bolus. To examine cardiac-driven pulsatility, we model CBF(*t*) with a 2nd-order Fourier model, which has previously been used for cardiac-gated measurements in ASL^13,17,27^. This model of CBF(*t*) is:

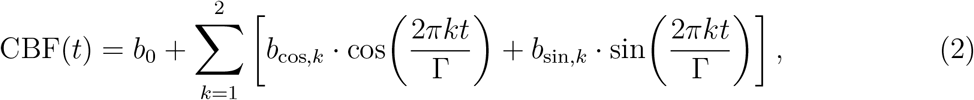

where *b*_0_ (= CBF_0_), *b*_cos,1_, *b*_sin,1_, *b*_cos,2_ and *b*_sin,2_ are the Fourier coefficients, and r represents the mean cardiac period (RR interval, or inverse heart rate). Evaluating the convolution in Equation 1 yields:

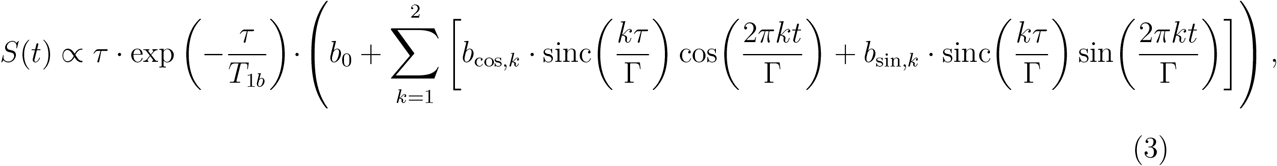

a smoothed version of the CBF(*t*) driving function with the coefficients modified by an order-dependent sinc(*kτ/*Γ) term, where 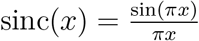.

The pulsatility of a given waveform *S* can be quantified via the pulsatility index (PI)^9,10,19,28^, which is given by:

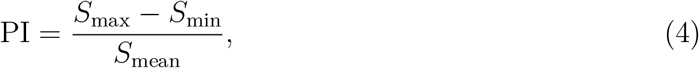

where *S*_max_ is the maximum of *S*(*t*), *S*_min_ is the minimum and *S*_mean_ is the mean.

By applying Equation 4 to the VSASL signal in Equation 3, a model for PI as a function of *τ* can be derived, with the full expression of PI(*τ*) given in Equation S.10 of Section S1. Although useful to examine, Equation S.10 is not an ideal platform for further analysis as it involves evaluating extrema of a second-order Fourier function (which do not simplify further) and is parametrized by CBF coefficients (whose values are generally not known *a priori*). However, in physiological flow waveforms, the fundamental (1st-order) frequency is typically the largest harmonic in the power spectrum^29^. By assuming 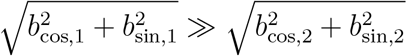 and neglecting the 2nd-order terms of *S*(*t*) (see Section S1 for mathematical justification of this approximation), Equation 4 can then be applied to the VSASL signal in Equation 3 to yield:

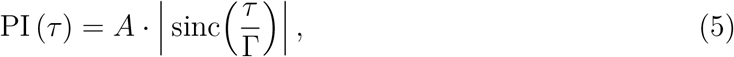

where 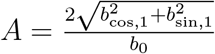 is a lumped fitting parameter. This simple expression describes a sinc-shaped dependence of PI on the ratio of the bolus duration *τ* (an adjustable VSASL scan parameter) to cardiac period Γ. Note that this form predicts that PI will vanish when *τ* = Γ, a feature that will be examined later with in vivo data.

### 3.2 Optimal Choice of Tau

We also determine the value of *τ* that maximizes the SNR of PI. In Section S3, we show that the SNR of the PI measurement is approximately proportional to the product of the SNR of the VSASL signal (∝*τ* · exp (−*τ*/*T*_1*b*_)) and the magnitude of PI (Equation 5):

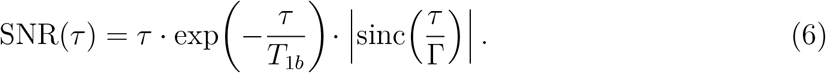

This expression is maximized at *τ*_opt_:

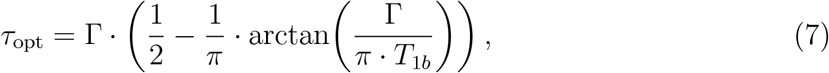

yielding an optimal value slightly below Γ*/*2. Figure 2A is a graphical demonstration of optimizing Equation 6 to obtain Equation 7. For an example cardiac period of Γ= 1, the optimal tau is computed as *τ*_opt_ = 0.437 s and indicated on the graph, and Figure 2B then plots Equation 7 to show *τ*_opt_ over a range of cardiac periods. In Figure 2A, we also note that the SNR(*τ*) curve is relatively flat around *τ* = *τ*_opt_. For example, the nearby point of SNR(Γ*/*2) = 0.219 (indicated by the black circle) is within 2% of SNR(*τ*_opt_) = 0.223. Figure 2C shows that the discrepancy between SNR(*τ*_opt_) and SNR(Γ*/*2) is small for a range of Γ (*<* 3% for typical Γ in [0.6, 1.2] s), suggesting that nearby *τ* values such as Γ*/*2 can also roughly optimize SNR with relatively little tradeoff.

**Figure 2:**
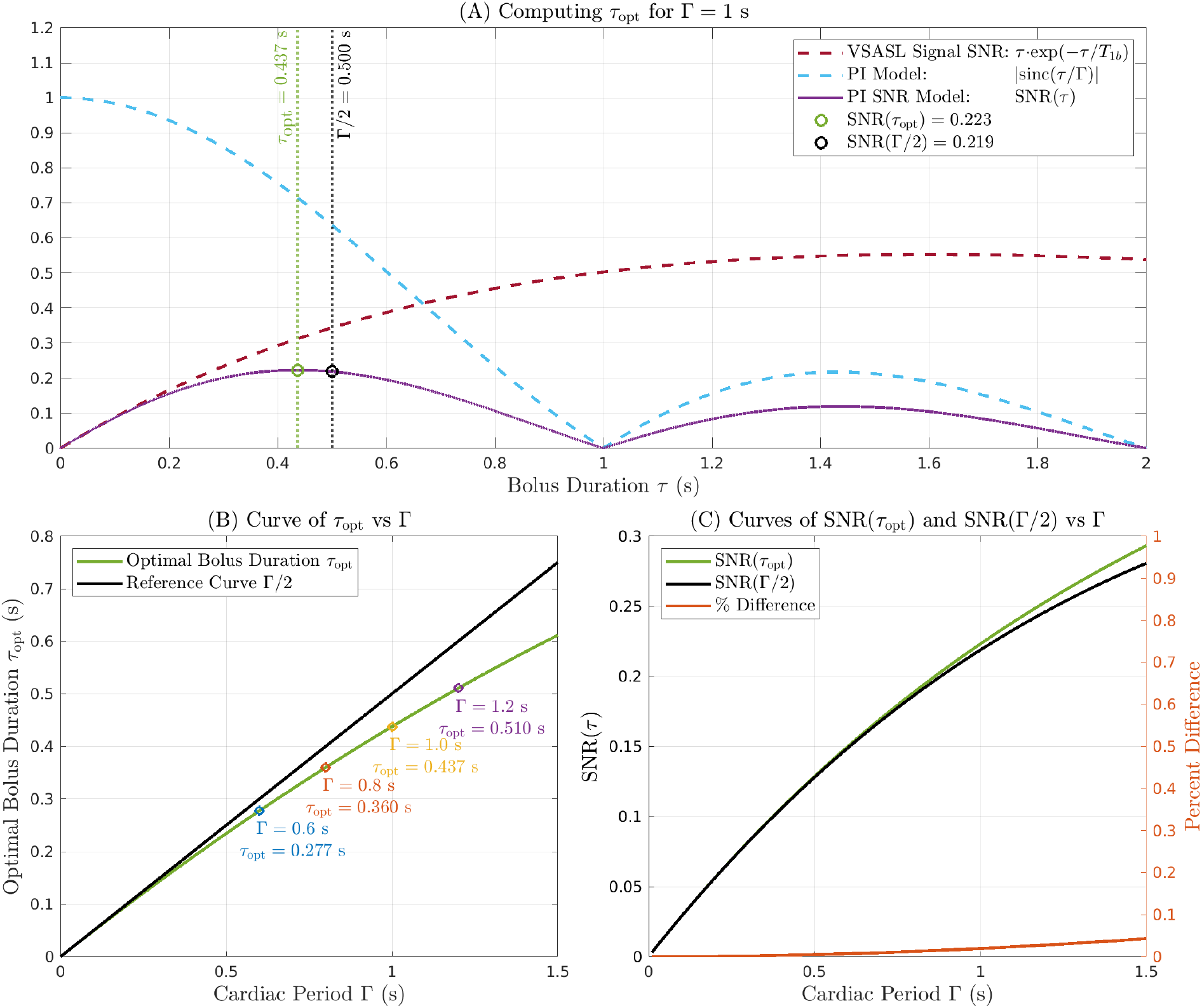
(A) Graphical representation of determining optimal tau *τ*_opt_ following Equations 6 and 7, for an example cardiac period Γ = 1 s. The red dashed line indicates the VSASL signal SNR model *τ* · exp (− *τ /T*_1*b*_). The blue dashed line indicates the PI sinc model of Equation 5. The purple solid line indicates the combined PI SNR model (Equation 6), which is optimized at *τ*_opt_ (computed via Equation 7), as indicated by the green circle. (B) A plot of *τ*_opt_ vs Γ per Equation 7. Several points are highlighted to indicate *τ*_opt_ for values of Γ in a typical physiological range. (C) A plot of SNR(*τ*_opt_) and SNR(Γ*/*2). The discrepancy between SNR(*τ*_opt_) and SNR(Γ*/*2) is shown to be fairly small (*<* 3% for r in [0.6, 1.2] s), suggesting that values near *τ*_opt_, such as Γ*/*2, could also yield near-optimal SNR.

A supplementary analysis on optimal *τ* is provided in Section S3, which presents the theory behind Equation 6 and the approximations used in its derivation. As additional support, the theoretical SNR model of Equation 6 is also compared with a reference SNR curve derived from Monte Carlo simulations. For those simulations, measurement noise was first added to simulated VSASL signals, distributions of PI measurements were computed, and then SNR was calculated from those PI distributions, with the process repeated over a range of *τ* values. As shown in the Section S3.3 results, Equation 6 shows excellent agreement with the simulated SNR curve, and *τ*_opt_ indeed yields close to the maximum SNR of the PI measurement.

## 4 Methods

### 4.1 Overview

A primary cohort of 7 healthy subjects (3 females and 4 males, aged 38.1 *±* 14.9 years), plus a secondary cohort of 10 relatively older healthy subjects (9 females and 1 male, aged 65.9 *±* 10.0 years), were enrolled in this study. The study was approved by the UCSD Institutional Review Board (IRB), and informed consent was obtained from all participants. The experiment consisted of a series of MRI scans, all acquired on a 3T MAGNETOM Prisma (Siemens, Erlangen, Germany) using a 64ch head/neck receiver coil. During the MRI scans, photoplethysmography (PPG) data were collected from the subject’s index finger using the built-in system sensor. These PPG data were used for retrospective cardiac gating.

### 4.2 MRI Acquisition

The scanning protocol for the primary cohort consisted of a localizer, a *T*_1_-weighted structural scan, and then a series of six VSASL scans: two at *τ* = 500 ms (to evaluate testretest repeatability near *τ*_opt_ per Equation 7), and then one each at progressively longer values (750/1000/1250/1500 ms) for a *τ* -stepping experiment to test for the predicted sinc-dependence described in Equation 5. All primary cohort subjects completed the full protocol, with a few exceptions noted in Table 2. The secondary cohort of 10 subjects underwent a short scanning protocol consisting of a *T*_1_-weighted structural scan and a single VSASL scan at *τ* = 500 ms, with all subjects completing this short protocol. *T*_1_-weighted structural scan parameters are provided in Section S4.

**Table 1:**
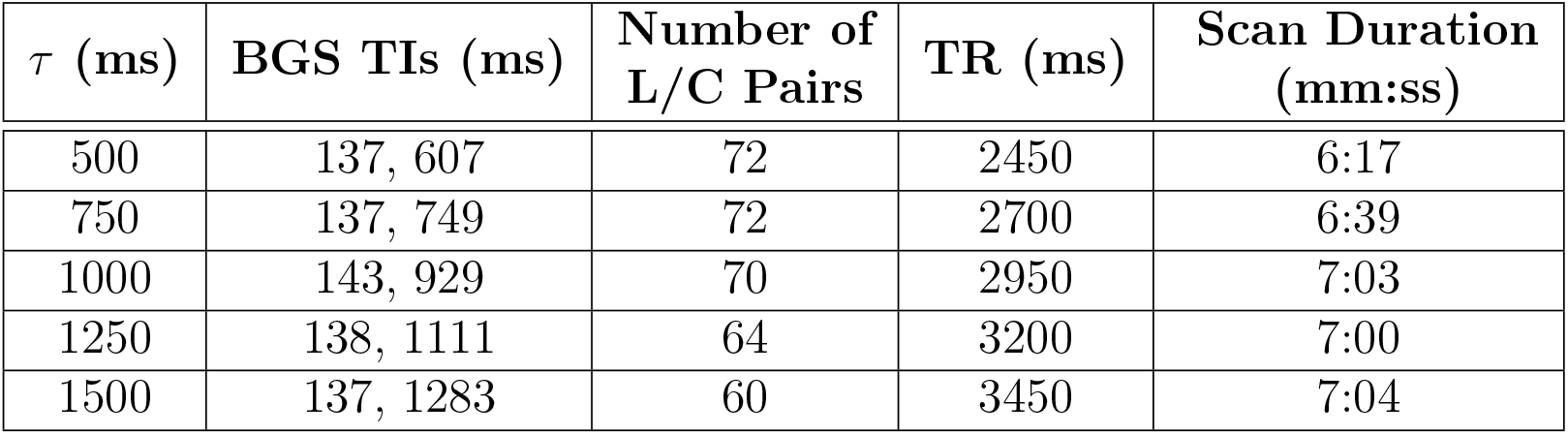
Table of VSASL scan parameters, indicating bolus duration *τ*, background suppression inversion times (BGS TIs), number of label/control pairs, repetition time *TR*, and total scan duration. All scans used *T*_*sat*_ = 1500 ms and *PLD* = 100 ms. Also, all VSASL scans contained one *M*_0_ (proton density-weighted) repetition at the beginning of the sequence with *TR* = 5000 ms. The BGS timings were configured via a MATLAB optimization routine to target complete suppression (0% signal) of GM and CSF tissue signals at 50 ms before the start of readout. This routine also considered *T*_2_-decay during the label-control module (LCM) and vascular crushing module (VCM) assuming an effective echo time (eTE) of 22 ms, and *T*_1,*GM*_ = 1300 ms, *T*_2,*GM*_ = 100 ms, *T*_1,*CSF*_ = 4000 ms and *T*_2,*CSF*_ = 2000 ms.

| $\tau$ (ms) | BGS TIs (ms) | Number of L/C Pairs | TR (ms) | Scan Duration (mm:ss) |
| --- | --- | --- | --- | --- |
| 500 | 137, 607 | 72 | 2450 | 6:17 |
| 750 | 137, 749 | 72 | 2700 | 6:39 |
| 1000 | 143, 929 | 70 | 2950 | 7:03 |
| 1250 | 138, 1111 | 64 | 3200 | 7:00 |
| 1500 | 137, 1283 | 60 | 3450 | 7:04 |

**Table 2:** Table of subject demographics and acquired data. Test-Retest refers to subjects who acquired two repeats of the *τ* = 500 ms scan. *τ* -stepping refers to subjects who completed the *τ* -stepping protocol consisting of scans at *τ* = 500*/*750*/*1000*/*1250*/*1500 ms. Only one *τ* = 500 ms scan was acquired on Subject 1 due to time constraints. This was rectified with longer scheduled scan sessions for all subsequent subjects. Both *τ* = 500 ms scans were acquired on Subject 3, but the first was excluded from analysis due to head motion. Subject 7 faced time constraints from late arrival, so only the two scans at *τ* = 500 ms were collected. Note: The genders were redacted and the ages were replaced by a range for the medRxiv submission.

| Subject | Gender | Age Range | Test-Retest | τ-Stepping |
| --- | --- | --- | --- | --- |
| 1 | - | 20-29 y.o. | - | x |
| 2 | - | 40-49 y.o. | x | x |
| 3 | - | 20-29 y.o. | - | x |
| 4 | - | 20-29 y.o. | x | x |
| 5 | - | 20-29 y.o. | x | x |
| 6 | - | 50-59 y.o. | x | x |
| 7 | - | 60-69 y.o. | x | - |

The VSASL pulse sequence consisted of a velocity-selective preparation module and an accelerated 3D gradient- and spin-echo (GRASE) readout. VSASL scans were configured for the five different bolus durations of *τ* = 500/750/1000/1250/1500 ms, with repetition time (TR) adjusted to accommodate the *τ* setting of a given scan. Other sequence timing parameters *T*_*sat*_ and post-labeling delay (PLD) were kept fixed across all scans at *T*_*sat*_ = 1500 ms and PLD = 100 ms, respectively. The PLD was kept to the minimum necessary to accommodate two spectrally-selective fat-saturation modules and two inferior saturation modules (to minimize CSF inflow effects) immediately prior to readout. Two background suppression (BGS) pulses were inserted for each scan between the LCM and the VCM. Additional sequence timing details are provided in Table 1.

An eight-segment 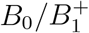 insensitive rotation (BIR-8) train^23,30^ was used for velocity-selective saturation for both the LCM and the VCM, with velocity weighting in the inferior-superior direction. The BIR-8 train was configured with a cutoff velocity of *v*_cut_ = 2 cm/s, with cutoff velocity defined as the first zero-crossing of the laminar flow response of the label condition^23^.

The 3D GRASE readout was configured as: matrix size = 56 *×* 56 *×* 24; voxel size = 4 *×* 4 *×* 6 mm^3^; FOV = 224 *×* 224 *×* 144 mm^3^; 10% Slice Oversampling; 6/8 Phase Partial Fourier; 2 *×* 2 GRAPPA Acceleration; Turbo Factor = 13; EPI Factor = 21; 1 segment (single-shot acquisition); flip angle = 120 degrees; TE = 12.62 ms.

### 4.3 Data Pre-Processing

For each subject, all VSASL scans were co-registered and motion-corrected using AFNI’s `3dvolreg` command^31^. The *T*_1_-weighted structural scans were processed using FSL’s `fsl_anat` command^32,33^, producing a whole-brain mask and a partial volume map of gray matter (GM). The GM partial volume map was thresholded at 0.8 (80%) to produce a GM mask. The brain and GM masks were nearest neighbor-resampled to the VSASL scan resolution using AFNI’s `3d_resample`^31^.

To minimize CSF contamination^23^, the VSASL labels and controls were pair-wise subtracted in MATLAB R2021A (Natick, MA) to form a perfusion-weighted time series, and then the median absolute deviation (MAD) was computed voxel-wise across the time dimension. To identify high-variance voxels contaminated by CSF, a whole-brain MAD threshold was defined at the 80th-percentile, along with a set of slice-specific MAD thresholds defined at the 80th-percentile within every brain slice. Voxels with MAD values above either the whole-brain or slice-specific thresholds were excluded from the GM mask, and the resulting mask served as the final GM ROI for the subsequent analysis.

Data were denoised using the iterative denoising method described in Power et al.^34^, with a few modifications: incorporating 2nd-order Fourier regressors (to preserve desired cardiac-driven fluctuations) and censoring volumes with outlier Γ values (e.g. due to finger motion). Supplementary details on denoising are available in Section S5.

### 4.4 Data Analysis

For each VSASL scan, the denoised data were spatially averaged within the GM ROI to produce time series for control and label measurements, respectively. A cardiac phase ϕ was assigned to every data point via PPG-based retrospective gating^15,27^. The control and label time series were separately fit to 2nd-order Fourier models based on prior work^27^. The VSASL signal was then obtained by subtracting the control and label Fourier coefficients to yield:

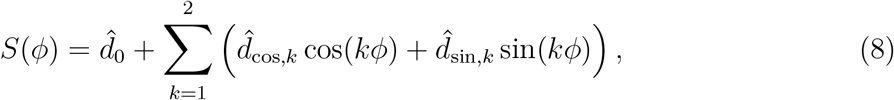

where 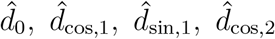 and 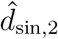 are the resultant Fourier coefficients of the signal *S*(*ϕ*). After obtaining *S*(*ϕ*), PI was quantified using Equation 4. The stability of the PI measurements was then assessed using a residuals permutation approach^35^. This procedure was repeated over 1000 iterations, and the resulting distribution of PI values was used to compute 95% confidence intervals. Additional details on the *S*(*ϕ*) computation and residuals permutation approach are provided in Section S2.

The measured PI values from the *τ* -stepping VSASL scans were fit to a version of Equation 5 modified to account for heart rate variability:

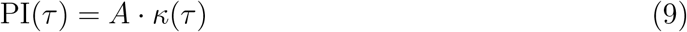

where *К* (*τ*) is the average of |sinc 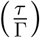| functions evaluated over all the Γ_*i*_’s observed during the scanning session:

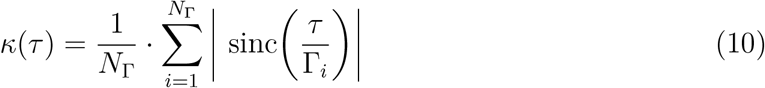

and *N*_Γ_ is the number of cardiac periods Γ observed. This effectively weights the model based on the distribution of Γ_*i*_’s observed during the scanning session. The model fits were evaluated using R^2^.

Using subjects with two runs of the *τ* = 500 ms scan, the test-retest repeatability of the GM ROI PI measurement at *τ* = 500 ms was assessed by using the Pearson correlation coefficient and intraclass correlation coefficient (ICC).

The association of PI with age across subjects was also assessed. Because PI depends on the ratio of *τ /*Γ, the comparison across subjects and conditions is facilitated by evaluating PI at a common reference ratio of *τ /*Γ = 0.5 (or equivalently, *τ* = Γ*/*2), which was chosen to minimize the extrapolation from the *τ* = 500 ms measurements (given that most observed cardiac periods were around Γ= 1000 ms). Based on Equation 9, we computed:

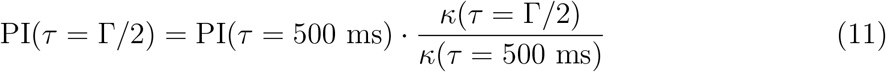

to yield a PI metric that was adjusted for individual cardiac periods and therefore comparable across subjects. For subjects with two *τ* = 500 ms scans, we performed the computation twice and averaged the PI(*τ* = Γ*/*2) values. Then, the relationship between PI(*τ* = Γ*/*2) vs age was assessed using the Pearson correlation coefficient.

## 5 Results

The fits of the subject data to the model in Equation 9 are shown in Figure 3, with R^2^ values ranging from 0.735 to 0.991 and minima occurring near *τ* = Γ as predicted by Equation 5. Notably, these subjects represent a diverse sample of cardiac periods, with values of Γ ranging from about 0.7 s to 1.1 s. Figure 4 serves as a complementary figure demonstrating the dependence of PI on *τ* for a representative subject (Subject 2) with a cardiac period around Γ = 1081 ms. In both the images and the curves, the pulsatility observed in the *τ* = 500 ms scans largely vanishes at *τ* = 1000 ms, where the image intensities and signal curves flatten out across the cardiac cycle. This was not unexpected given that the value of *τ* = 1000 ms is close to r, where the sinc 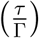 term of Equation 5 evaluates to 0.

**Figure 3:**
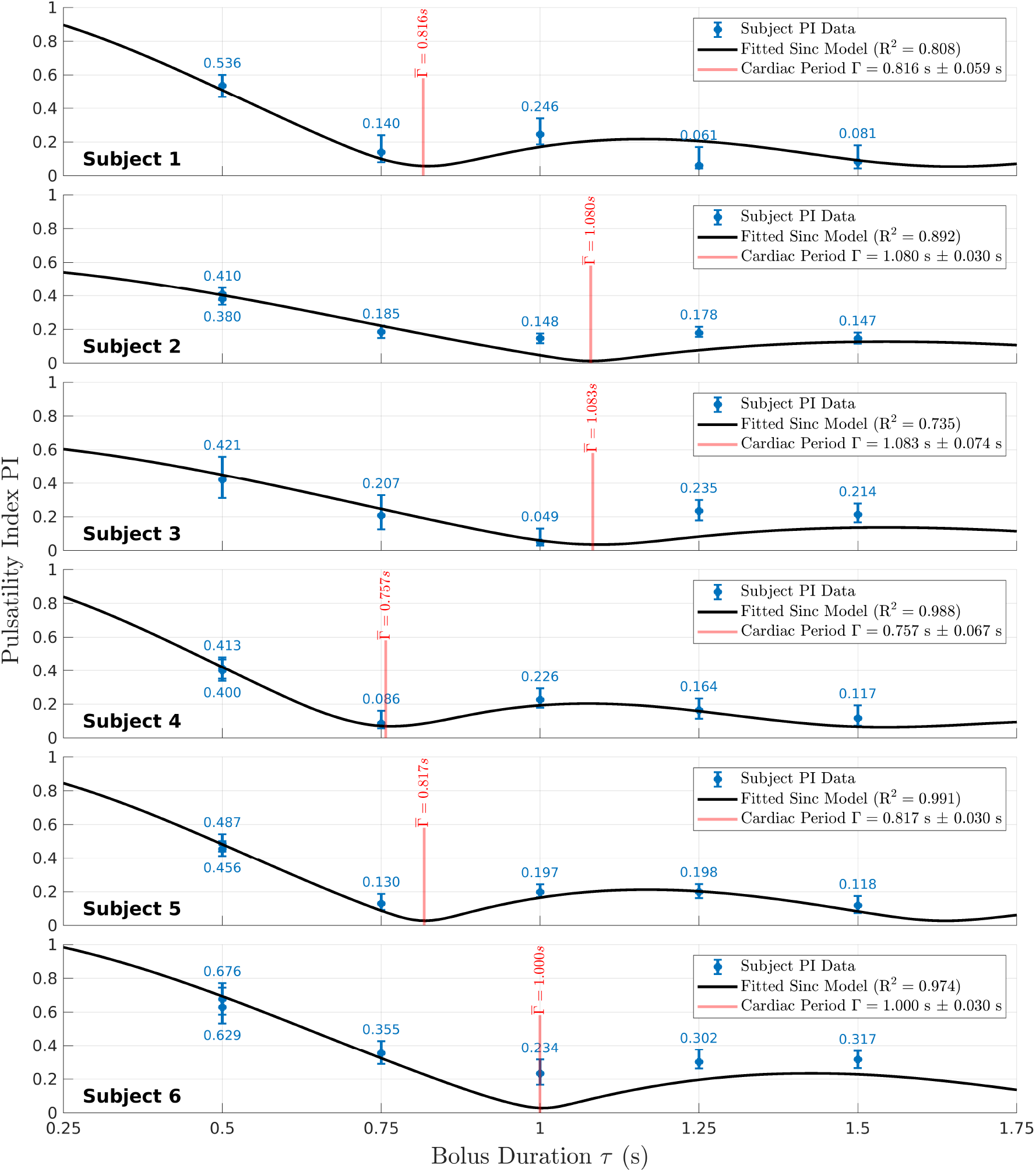
Fits of the modified sinc model (Equation 9) to the PI values measured from the *τ* -stepping protocol on each subject. The blue data points indicate the PI measured at each *τ* scan, with the black curve representing the best fit modified sinc model of Equation 9. The model fits the data quite well across a range of heart rates represented by these subjects. We consistently see PI reach a minimum around *τ* = Γ as predicted by Equation 5.

**Figure 4:**
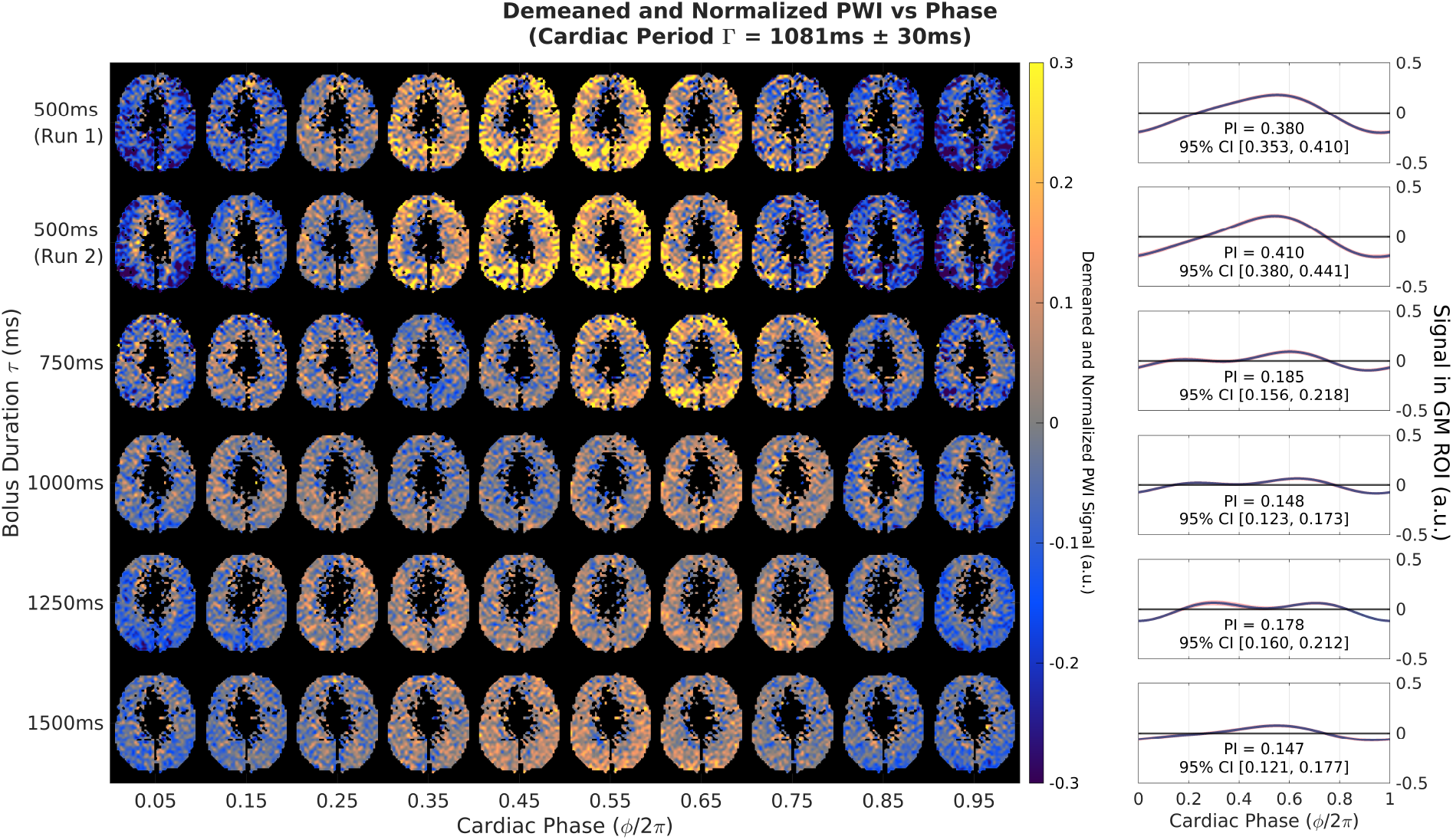
The left-hand montage shows the demeaned and normalized perfusion-weighted images (PWI) for a select slice of Subject 2, with cardiac phase *ϕ/*2*π* indicated along the x-axis and *τ* values for each scan indicated along the y-axis. To produce the maps on the left-hand side, the data are fit on a voxel-wise basis to obtain *S*(*ϕ*) curves for every voxel, then voxel-wise demeaned, and finally normalized by the scalar GM ROI mean value. The voxel intensities thus represent the relative fluctuation amplitude around each voxel’s mean compared to the baseline GM value. The voxels in the middle of the slices were masked due to ventricular location and CSF contamination (see Section 4.3). The accompanying line plots on the right-hand side show the GM ROI-averaged signal *S*(*ϕ*). The red shaded regions represents the 95% confidence interval at each value of *ϕ/*2*π*, derived from the set of *S*(*ϕ*) curves resulting from the residuals permutation approach. The text indicates the PI value, along with its 95% confidence interval. Note that for better visual comparison, the curves (and images) were phase shifted so that the maximum amplitude roughly aligns across the scans (rows) in this figure.

Figure 5 demonstrates the computation of a voxel-wise PI map using data from Subject 2 (corresponding to the same underlying data used in the first row of Figure 4). Following the same analysis procedures described above in Methods for the GM ROI signal, the VSASL signal *S*(*ϕ*) was computed on a voxel-wise basis, followed by a voxel-wise PI calculation via Equation 4.

**Figure 5:**
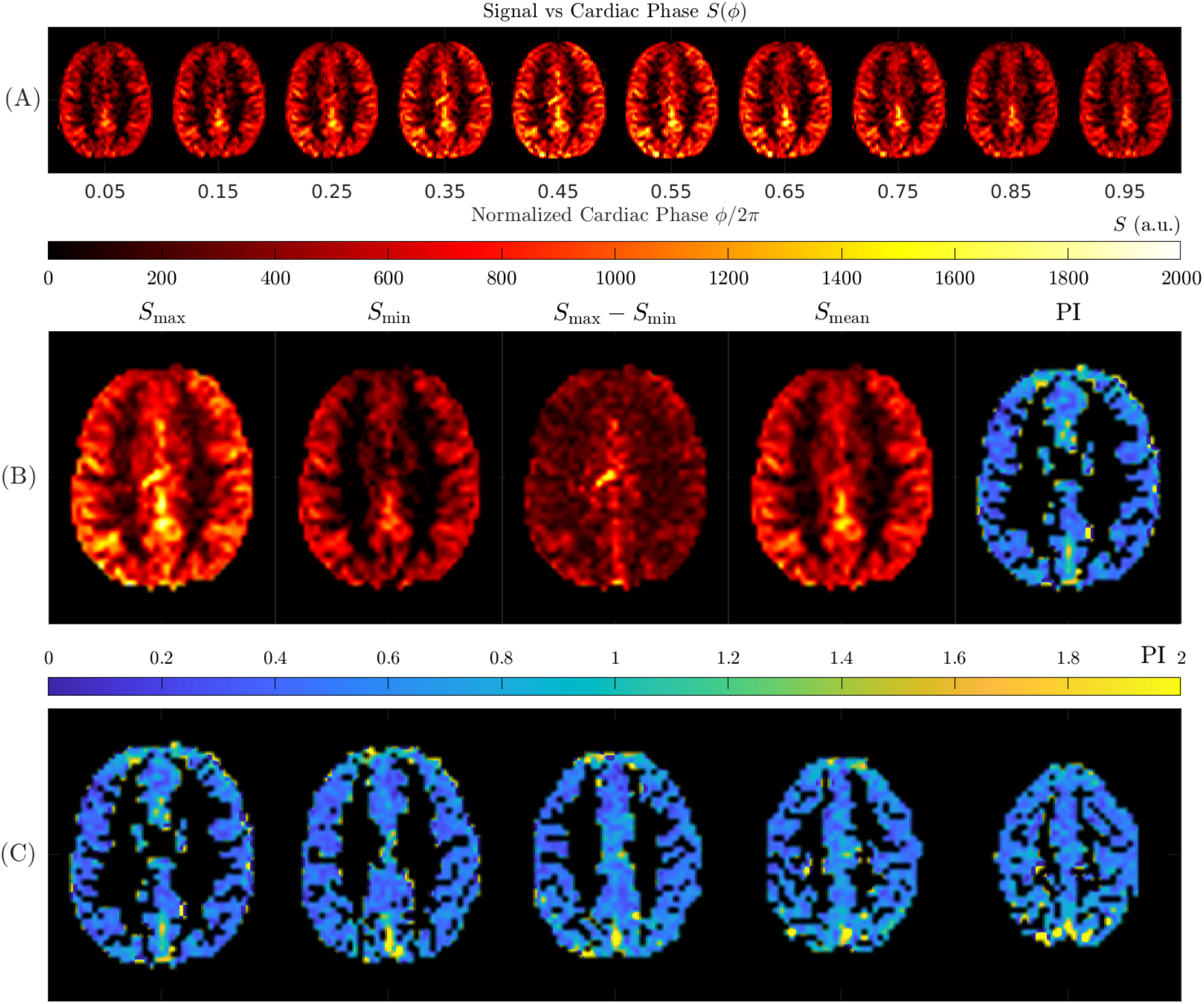
Demonstration of a voxel-wise PI map using Subject 2 data, *τ* = 500 ms, run 1. (A) Perfusion signal as a function of cardiac phase *S*(*ϕ*), computed as in Section 4.4 on a voxel-wise basis. Note that as compared to the demeaned maps shown in the top row of 4, the voxel-wise mean is preserved in these maps. (B) Individual components of the Equation 4 formula corresponding to *S*_max_, *S*_min_, *S*_max_ − *S*_min_ (the numerator of the PI formula), *S*_mean_ (the denominator of the PI formula), and the computed PI map. (C) Select slices of the same PI map. Note that in (B) and (C), the PI slices are masked in order to focus on gray matter voxels.

Figure 6 shows the repeatability of PI at *τ* = 500 ms across subjects. All data points lie close to the line of unity, with a high ICC (ICC = 0.960, *p <* 0.001) and correlation coefficient (*r* = 0.986, *p* = 0.002). A representative visual example of this repeatability is demonstrated in Figure 4 for Subject 2, where the top two rows show the repeats at *τ* = 500 ms. The appearances of the perfusion maps are nearly identical across the cardiac cycle. The morphologies of the GM ROI-averaged signal curves (Figure 4, right) are also nearly identical, with consistent shape and amplitude in both runs.

**Figure 6:**
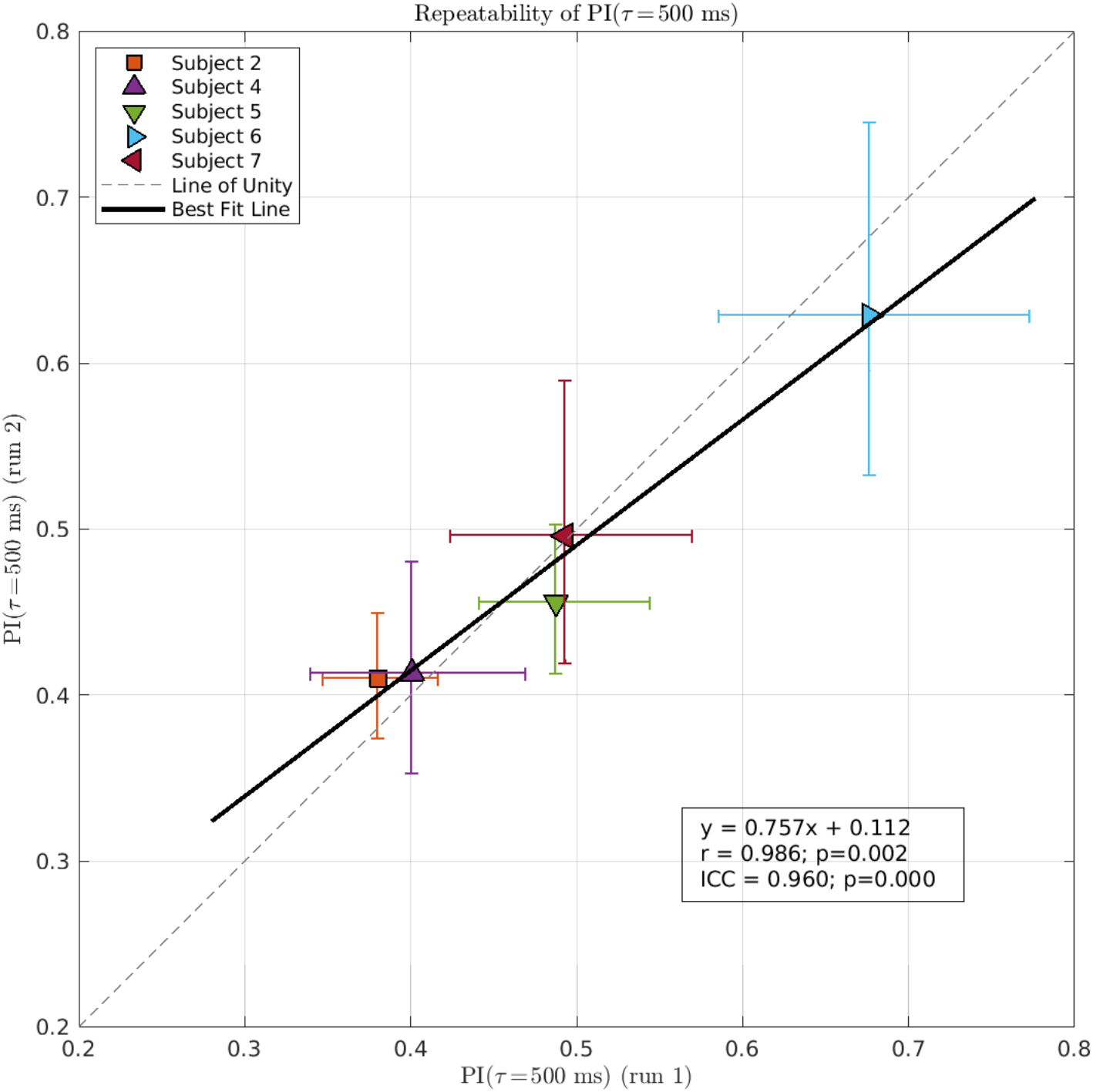
Repeatability of PI(*τ* = 500 ms) across subjects. The x-axis denotes the PI of run 1, and the y-axis denotes the PI of run 2. The data points are the measured values from the 5 subjects with repeats of the *τ* = 500 ms scan. The error bars indicate the 95% confidence intervals of the values. The values follow the line of unity closely, indicating excellent test-repeat repeatability. This is further supported by the high Pearson correlation coefficient *r* = 0.986 (*p* = 0.002) and ICC = 0.960 (*p<* 0.001).

Figure 7 shows a significant positive correlation between PI(*τ* =−*/*2) and age (*r* = 0.554, *p* = 0.021), showing the potential of this method to explore an association with age.

**Figure 7:**
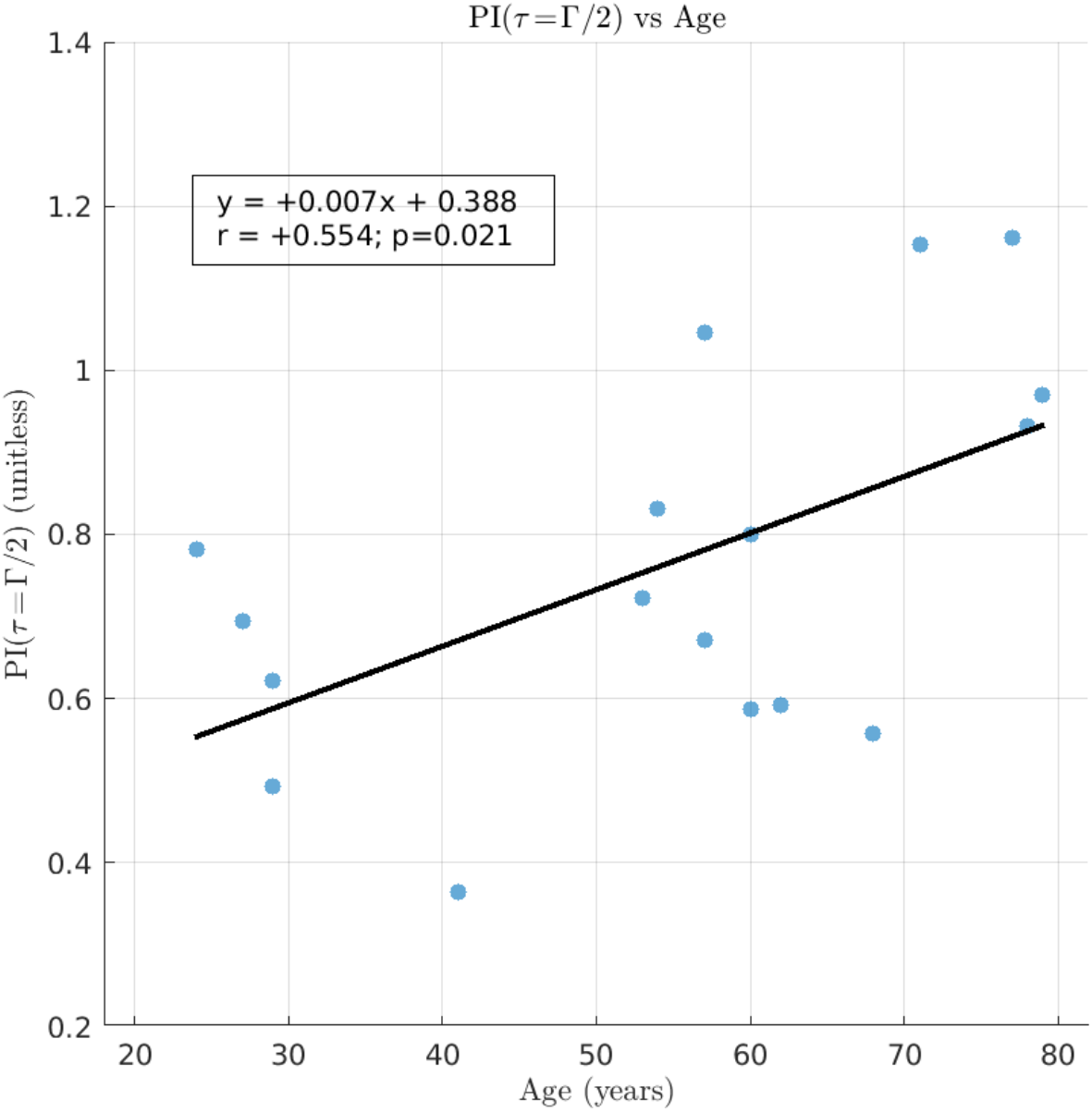
Plot of PI(*τ* = Γ*/*2) vs age, showing a positive correlation between PI(*τ* = Γ*/*2) and age (*r* = 0.554, *p* = 0.021).

## 6 Discussion

In this study, we have presented MVP-VSASL, a novel approach for measuring cerebral microvascular pulsatility by leveraging the microvascular specificity of VSASL and retrospectively gating the VSASL signal. A key innovation of our study is the simple theoretical model for VSASL signal pulsatility PI given in Equation 5, which provides an explicit form for PI as a function of bolus duration *τ* and cardiac period Γ. This sinc model is validated by its excellent fits to in vivo data from 6 subjects representing a broad range of cardiac periods. Furthermore, the model predicts a minimum in PI near *τ* = Γ, which is empirically observed in all subjects. This feature can be explained intuitively: A VSASL bolus of duration of *τ* = Γ integrates over one whole period of the periodic CBF signal, regardless of the position of the bolus in the cardiac cycle. This results in minimal fluctuation of the gated VSASL signal and thus a minimal value of PI.

Using this sinc model of Equation 5, we also derived a theoretically optimal value of *τ* in Equation 7 to maximize the SNR of the pulsatility measurement. While the optimal value *τ*_opt_ is slightly below Γ*/*2 based on theory, choosing *τ* = Γ*/*2 represents an SNR tradeoff of less than 3% for typical cardiac periods in the range of [0.6, 1.2] s. In this study, we used a pre-defined set of *τ* values in order to keep the scan configurations consistent across subjects. Using a fixed value of *τ* = 500 ms for the repeatability and age-related scans in this study corresponded to a value of *τ* that was reasonably close to the sample (both cohorts combined) mean Γ*/*2 value of 460 ms (range 380 ms to 570 ms) and yielded predicted SNR values that were on average within 7% of the optimal SNR. Future work will be useful for better determining the tradeoffs between using a fixed *τ* (e.g. *τ* = 500 ms) or selecting a scan-specific *τ* = Γ*/*2 (which requires adjusting the scan timing parameters on a per-scan basis).

We also show the potential of the metric PI(*τ* = Γ*/*2) for detecting physiological differences across subjects, as the positive association of PI(*τ* = Γ*/*2) with age (Figure 7) agrees well with previous studies finding higher flow pulsatility (as measured by PC-MRI) in older vs younger subjects^8,10,22^. While future work is needed to validate this association in larger populations, the agreement with prior literature supports the validity of the proposed VSASL approach.

### 6.1 Comparison with other techniques

The strength of MVP-VSASL is the ability to measure pulsatility in the microvasculature with vessel diameters around 50 µm. This is notably different from the majority of current pulsatility approaches, which focus on larger arteries feeding the brain. This focus on large vessels is a common feature across different modalities and techniques (e.g. 2D PC-MRI^8,9^, 4D flow^11,36^, doppler ultrasound^4,12^, or ASL^13–15,17^) and different measures of pulsatility (e.g. pulsatility of blood volume^13–15,17^, vessel distension^16^, flow velocity^12^, or flow volume rate^4,8,36^). For blood volume pulsatility, the exceptions to our knowledge are the dynamic ASL approach by Yan et al.^14^, where significant differences in blood volume between systole and diastole were found in the small arteries and arterioles, and the 7T VASO approach by Guo et al.^37^, which can measure relative blood volume over the cardiac cycle to estimate a vascular compliance index.

To our knowledge, the only other method that measures flow pulsatility in the microvasculature is PC-MRI at 7T^19–22^, which targets cerebral perforating arteries with diameters below 300 µm and flow velocities as low as 0.5-1.0 cm/s. This approach involves resolving and measuring the flow through individual vessels, averaging the flow curves across vessels, and computing pulsatility on the averaged flow signal. For an approximate comparison, the 7T PC-MRI microvascular pulsatility values in the basal ganglia (a deep gray matter region) were reported to be around 0.55-0.65 for young subjects (mean age around 25 years)^22^, and around 0.7-1.0 for older subjects (mean age around 60-65 years)^21,22^. Using an age cutoff of 45 years, our PI(*τ* = Γ*/*2) values in cortical GM are 0.591 *±* 0.165 for younger subjects (n = 5, age 30.0 *±* 6.48 years) and 0.835 *±* 0.217 for older subjects (n = 12, age 64.7 *±* 9.56 years), which agree well with the 7T PC-MRI range of values. While the pulsatility values from 7T PC-MRI serve as a useful reference point, a direct comparison of their pulsatility values to those of the current study should be interpreted with caution due to (1) the different ROIs used between studies, and (2) the effect of bolus duration in MVP-VSASL. Specifically, the 7T PC-MRI studies have focused on the centrum semiovale (a white matter region) and basal ganglia, whereas the current VSASL study focuses on cortical GM. Future work assessing the same ROIs and exploring an appropriate adjustment for the *τ* -dependent sinc factor would be useful for a more direct comparison between MVP-VSASL and the 7T PC-MRI approach.

Our VSASL approach builds upon the prior body of work examining cardiac-driven fluctuations^26,38–41^ and pulsatility^13–15,17^ of the ASL signal. While the majority of this literature focused on spatially-selective ASL techniques^24^, Franklin et al.^26^ examined VSASL signal fluctuations weighted by macrovascular blood flow and volume (due to the absence of a VCM), and did not explicitly compute a pulsatility index. Other ASL-based pulsatility measurements have used spatially-selective ASL approaches^24^ to assess cerebral blood volume (CBV) pulsatility and vascular compliance in mainly large arteries^13–15,17^, whereas MVP-VSASL focused on a flow pulsatility measurement in the microvasculature. For flow-weighted ASL signals, some studies recognized that selecting a bolus duration equal to the cardiac period (or a multiple) could mitigate cardiac-driven fluctuations^27,38^, which we noted (both theoretically and empirically) as well. However, we are the first to our knowledge to derive a model (Equation 5) explicitly describing the dependence of cardiac fluctuations on *τ* and Γ, providing a useful theoretical extension to predict the magnitude of such signal fluctuations.

### 6.2 Clinical Considerations

An accurate and reliable measurement of microvascular pulsatility can be valuable for clarifying the mechanisms connecting pulsatility, microvascular damage, and cognitive decline in various diseases. Previous studies have associated decreased performance over a range of cognitive domains with pulsatility in large cerebral arteries^4,9,12^, but these measurements remain distant from the microvasculature, where the damage linked to various cognitive disorders is likely occurring^4^. In this regard, microvascular pulsatility measurements may be more suitable for assessing the relevant environment. Furthermore, since vascular risk factors such as microvascular pulsatility may reflect early and potentially modifiable variables in disease progression^5–7,18,42^, MVP-VSASL could also help assess cognitive risk before structural changes and clinical symptoms of cognitive impairment arise.

Because large arteries each supply pulsatile blood to extensive vascular territories that include many regions of the brain, the large-artery pulsatility indices are inherently limited in their spatial specificity when interpreting their impact on specific brain regions. In comparison, pulsatility measurements in the microvasculature, being much more distal along the arterial tree and embedded in the tissue they are supplying, can potentially be used to probe specific areas implicated in disease pathogenesis (e.g. the parietal lobe and hippocampus for Alzheimer’s disease). MVP-VSASL, with its whole-brain coverage and the ability to simultaneously generate microvascular signals across the entire brain, offers a unique potential to facilitate such regional assessments. By computing PI on a voxel-wise basis, we have demonstrated the feasibility of generating a voxel-wise pulsatility map as shown in Figure 5. To our knowledge, this is the first demonstration of a microvascular pulsatility map, and the robustness and applicability of the approach for regional pulsatility assessments will be explored in future work.

Importantly, our technique has several practical features that may ease translation, including the ability to perform at 3T, a simple prescription with whole-brain coverage, and a clinical scan time of around 6 minutes. These features make MVP-VSASL an attractive option for integration into standard clinical and research brain MRI protocols.

### 6.3 Technical Considerations

In this study, we used the standard VSASL cutoff velocity of *v*_cut_ = 2 cm/s^23^, which defines the VSASL bolus in the microvascular regime. However, the value of *v*_cut_ can be adjusted to target other segments of the arterial network. Increasing *v*_cut_ shifts the VSASL bolus-defining location more upstream into larger vessels, whereas decreasing *v*_cut_ shifts the labeling region further distally toward the capillaries. Varying *v*_cut_ could thus be useful for evaluating pulsatility along the arterial tree using a single technique (by adjusting *v*_cut_) and assessing metrics such as the vascular dampening factor^8^. Of note, decreasing *v*_cut_ requires higher gradient strengths, which can exacerbate known technical issues like eddy currents, diffusion effects, and CSF contamination, but further improvements to the VSASL pulse train could make this exploration more feasible in the future^23^.

Prior studies on VSASL have explored its potential to measure venous flow^43,44^. The current analyses could also be applied to measure venous flow pulsatility, which has already received some attention using other techniques^11^.

A potential challenge of our approach is CSF contamination of the VSASL signal. This is an existing issue with VSASL in general, as CSF is difficult to suppress through standard background suppression techniques^23^. When using VSASL for quantifying mean cerebral blood flow, this manifests as an overestimation of mean perfusion in CSF-contaminated regions. For our study, CSF contamination may also affect signal fluctuations as CSF can also pulsate with the cardiac cycle^45^. Our pre-processing attempted to mitigate CSF effects as much as possible by aggressively excluding contaminated areas from the GM masks, but we acknowledge that it is not possible to completely eliminate CSF partial volume effects. Furthermore, these effects are challenging to remove through modeling due to the complex and heterogeneous nature of CSF flow. However, as VSASL sequences continue to improve, our approach can benefit from future advances related to CSF suppression^46,47^.

Finally, we used a standard BIR-8 train for VSASL labeling in this proof-of-principle study. The BIR-8 train is a velocity-selective saturation (VSS) technique noted for its robustness to 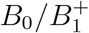 inhomogeneity and eddy currents, making it well suited for this initial study. Velocity-selective inversion (VSI) approaches that provide increased SNR^48^ were attempted, but resulted in too many artifacts particularly at lower bolus duration values of *τ*. While the dynamic phase cycling method mitigates many of these artifacts^49^, the resulting four-fold decrease in temporal resolution renders the retrospective gating method impractical within a reasonable imaging time. More robust VSI techniques better suited for our pulsatility method are currently under investigation^50^.

## 7 Conclusion

We have introduced MVP-VSASL, a novel, practical and non-invasive approach of using VSASL to measure pulsatility in the microvasculature, including a theoretical framework relating the pulsatile VSASL signal to the pulsatility index. This technique may be used to probe the mechanisms underlying cognitive disease, monitor disease progression, and evaluate patient responses to therapy.

## Data Availability

All data produced in the present study are available upon reasonable request to the authors.

## 8 Acknowledgments

The authors would like to thank Maria Bordyug, Lauren C. Edwards, Alin Alshaheri Durazo, Amanda I. Gonzalez, and Mary Ellen Garcia for their assistance in collecting the data.

## Supporting Information

### S1 Evaluating Sinc Model Approximation

In this section, we evaluate the error associated with the sinc model of pulsatility (Equation 5), whose derivation involves neglecting the 2nd-order Fourier terms. To evaluate the error, an exact expression for PI(*τ*) of a 2nd-order Fourier signal is derived. Then the exact PI(*τ*) curves (the reference) are computed over a range of *τ* values, and Equation 5 (the model) is fit to the reference to evaluate the model error.

#### S1.1 Theory

To provide an exact expression of PI, we start with the 2nd-order Fourier model for *S*(*t*) given in Equation 3, with the summation expanded out for clarity in this section. Note the *T*_1_ exponential decay factor is excluded for simplicity, as it cancels out in the final PI calculation.

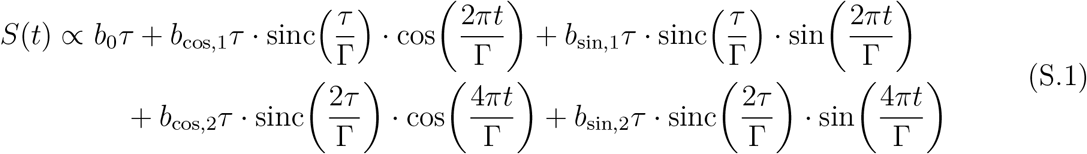

By combining the cos(*·*) and sin(*·*) terms of the same order, the equation is rewritten as:

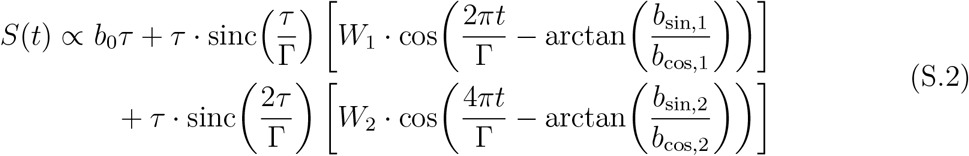

where

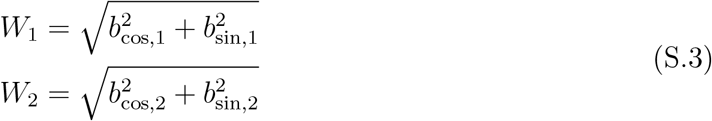

represent the magnitudes of the 1st and 2nd-order Fourier components, respectively. To compute PI, we can apply Equation 4 (reproduced below):

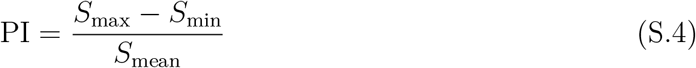

by evaluating the individual terms of the formula. The mean term *S*_mean_ is simply the constant term:

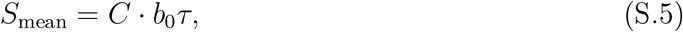

where *C* is the proportionality constant (which accounts for the switch from the ∝ symbol in Equation S.2 to the = symbol in Equation S.5). The max term *S*_max_ is computed by taking the maximum of *S*(*t*):

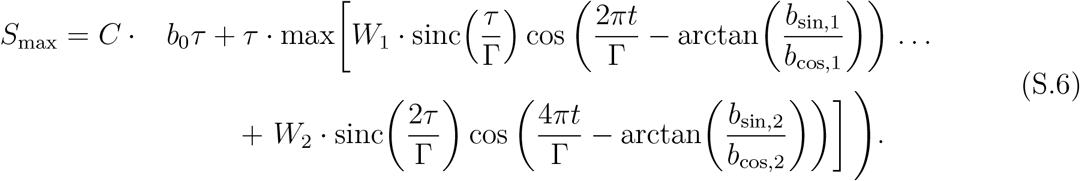

Because the max[*·*] operation is insensitive to phase shifts, *S*_max_ can be further rewritten as:

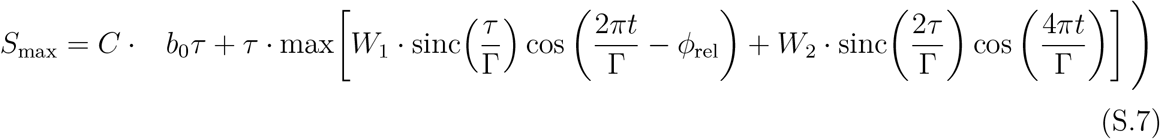

where *ϕ*_el_ is defined as:

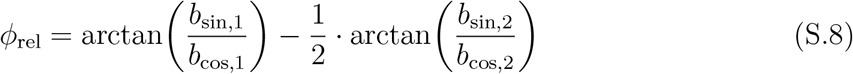

and represents the relative phase shift between the 1st- and 2nd-order sinusoids. Following the same logic, the min term *S*_min_ can be represented in a similar way:

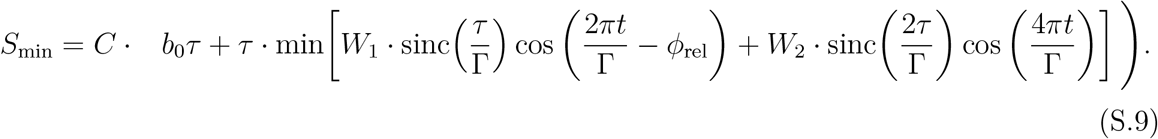

By substituting *S*_mean_, *S*_max_ and *S*_min_ into Equation 4, we obtain the following exact expression for PI(*τ*):

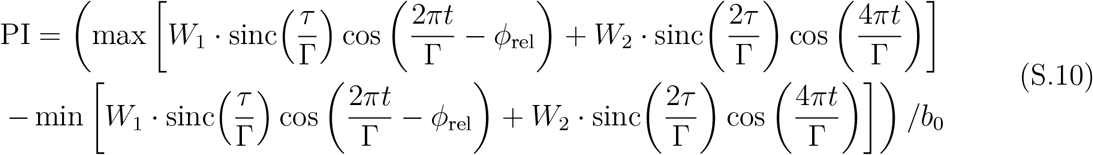

This is a formula for PI that depends on the scaling terms *b*_0_, *W*_1_, *W*_2_ and a relative phase shift between 1st- and 2nd-order cosines given by *ϕ*_rel_. Note that if the 2nd-order component is neglected by setting *W*_2_ = 0, then the formula simplifies down to the sinc model stated in Equation 5 of the main text and reproduced below for convenience:

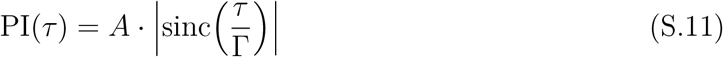

where 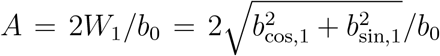 is the lumped fitting parameter described in the main text.

#### S1.2 Methods

To evaluate the errors associated with the sinc model approximation, we: (1) computed exact PI(*τ*) curves (Equation S.10), (2) fit the sinc model (Equation 5) to those exact curves, and (3) assessed the error of those fits.

To produce the exact PI(*τ*) curves, the constants (*b*_0_, *W*_1_, *W*_2_) were set at physiologically reasonable values based on our empirical measurements and on prior literature. First, *b*_0_ = 1 and *W*_1_ = 0.5 were chosen to ensure the PI(*τ*) curves have values comparable to in vivo data measurements at *τ* = 500*/*750*/*1000*/*1250*/*1500 ms. For example, this choice yields a value of PI(*τ* = Γ*/*2) = 0.636, which is in the middle of the range of values shown in Figure 7. While the exact *b*_0_ and *W*_1_ values are arbitrary, their ratio controls the vertical scaling factor 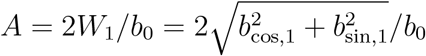 in the Equation 5 sinc model, which roughly scales the exact PI(*τ*) curves as well. To choose *W*_2_, prior estimates of blood flow power spectra in the internal carotid artery (ICA) were used to set the power ratio of the 1st-vs 2nd-order components 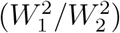 as 5:1^1^, which results in 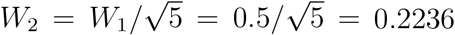. (Note that the power ratio in the microvasculature may even be higher than 5:1, since arterial compliance generally behaves like a low-pass filter on flow waveforms. This would comparatively attenuate the 2nd-order power as blood travels from the ICA to the microvasculature and result in an even smaller relative value of *W*_2_.) To summarize, the constants are set at *b*_0_ = 1, *W*_1_ = 0.5 and *W*_2_ = 0.2236.

A cardiac period of Γ = 1 s was set based on the range observed in subjects (approximately [0.7, 1.1] s). Bolus duration *τ* was varied over *τ* ∈ [0.400, 1.600] s in steps of 0.001 s. The remaining unconstrained parameter of *ϕ*_rel_ (the relative phase shift between 1st- and 2nd-order components) was varied between 0 to 2*π* in steps of 2*π τ* 0.001.

For each combination of *ϕ*_rel_ and *τ*, the exact value of PI(*τ*) (Equation S.10) was computed by numerically evaluating its terms over *t* ∈ [0, Γ] in steps of 0.0001 s. This resulted in curves of PI(*τ*) for every value of *ϕ*_rel_. Finally, for each value of *ϕ* _rel_, the sinc model (Equation 5) was fit to the exact PI values at *τ* = 500*/*750*/*1000*/*1250*/*1500 ms, which correspond to the *τ* values used in the *τ* -stepping experiment.

Since the derivation of the sinc model (Equation 5) involves neglecting 2nd-order terms of the signal curves, a natural question is whether we could simply start with a 1st-order Fourier model of the signal *S*, which would obviate the need for any approximation step for deriving the PI(*τ*) expression. As a complementary empirical analysis, we also re-analyzed the *τ* - stepping data (Figure 3 of the main text) by using a 1st-order Fourier model to describe *S*(*ϕ*), and compared the results to those of the 2nd-order approach. The same procedures outlined in Section 4.4 were followed, with the exception of using a 1st-order Fourier model to fit the control and label data of each individual scan. After fitting the *τ* -stepping data to the sinc model (Equation 9), the resulting values of *A* (i.e. the scaling factor of the sinc model) and R^2^ of the fits were also compared between the 2nd-vs 1st-order approaches using a paired Wilcoxon signed rank test.

#### S1.3 Results

The results of fitting the sinc model (Equation 5) to the exact PI values at *τ* = 500*/*750*/*1000*/* 1250*/*1500 ms are shown in Figure S1, with Figure S1A showing the best case fit based on R^2^ (over values of *ϕ*_rel_), Figure S1B showing the worst case fit, and Figure S1C the error across the entire space of *ϕ*_rel_. There is very little error across the entire space of *τ* and *ϕ*_rel_, supporting the validity of the sinc model (Equation 5) for describing pulsatility as a function of *τ*.

Figure S2 shows the results of the empirical *τ* -stepping analysis comparison. As shown in the insets of Figure S2A and Figure S2B, the main qualitative difference between the 2nd-vs 1st-order approaches is that the 1st-order model of *S*(*ϕ*) has difficulty describing relatively sharper peaks (which are captured by the 2nd-order approach), an effect most clearly shown for the *τ* = 500 ms scans. As a result, the 1st-order approach tends to underestimate PI for each scan, which can be seen in the individual subject examples (i.e. the orange data points being consistently lower than the blue ones). This effect is also summarized in Figure S2C, which shows a scatter plot of the PI values. The x-axis indicates the PI values resulting from the 1st-order approach, and the y-axis indicating the PI values resulting from the 2nd-order approach. Almost all points lie above the line of unity, indicating consistently higher PI values with the 2nd-order approach. However, the ability of the sinc model to describe the PI(*τ*) data seems to remain generally strong regardless of the 2nd-vs 1st-order approach taken to analyze the data for each individual *τ* scan. As shown in Figure S2D, the R^2^ values of the fits are all around 0.75 or higher (with one outlier exception in Subject 3 for the 1st-order approach), and the Wilcoxon signed rank test shows no statistically significant difference between the R^2^ values (*p* = 0.094). In summary, while using the 1st-order approach of *S*(*ϕ*) seems to underestimate the PI values, the sinc model still appears to describe the PI(*τ*) shape well with either approach in these comparisons.

### S2 Computing VSASL Signal and Pulsatility

This section provides supporting details for Section 4.4 of the main text, and describes the computation of the VSASL signal *S*(*ϕ*) as a function of cardiac phase and the subsequent computations of PI and confidence intervals.

The computation of *S*(*ϕ*) involves (1) retrospective gating to map control and label data (acquired in time) to cardiac phase space, (2) fitting 2nd-order Fourier models to control and label data points, and (3) subtracting the fits to obtain *S*(*ϕ*). We start with (*N ×* 1) control and label measurement vectors:

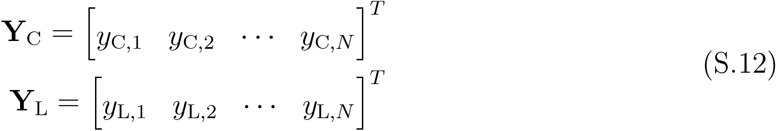

where *y*_C,*i*_ and *y*_L,*i*_ represent the *i*th control and label measurements, respectively, and *N* is the number of control/label pairs acquired from the scan. These measurement vectors can represent an ROI-averaged signal (as in the GM ROI-averaged signal used in Section 4.3) or the values from single voxels (as was used to produce the voxelwise PI map in Figure 5).

Next, a cardiac phase *ϕ* was assigned to every label and control data point by retrospectively gating on the photoplethysmography (PPG) waveform trace. First the peaks of the PPG waveforms were detected. Then, a cardiac phase *ϕ*_C,*i*_ or *ϕ*_L,*i*_ was assigned to every control and label volume, respectively, using:

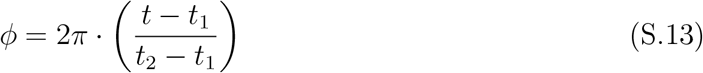

where *t* is set to *t*_C,*i*_ or *t*_L,*i*_ (defined as the center of the labeling/control period with width *τ*) for each volume, *t*_1_ is the preceding cardiac peak, and *t*_2_ is the subsequent cardiac peak^2^. For a given data point, time *t*_C,*i*_ or *t*_L,*i*_ was computed by subtracting *PLD* + *τ /*2 from the start time of the readout. The phase values were concatenated into (*N ×* 1) vectors for the controls and labels:

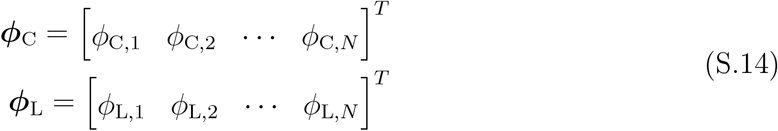

where *ϕ*_C,*i*_ and *ϕ*_L,*i*_ represent the cardiac phase of the *i*th control and label measurements, respectively. Then (*N ×* 5) 2nd-order Fourier design matrices were constructed as:

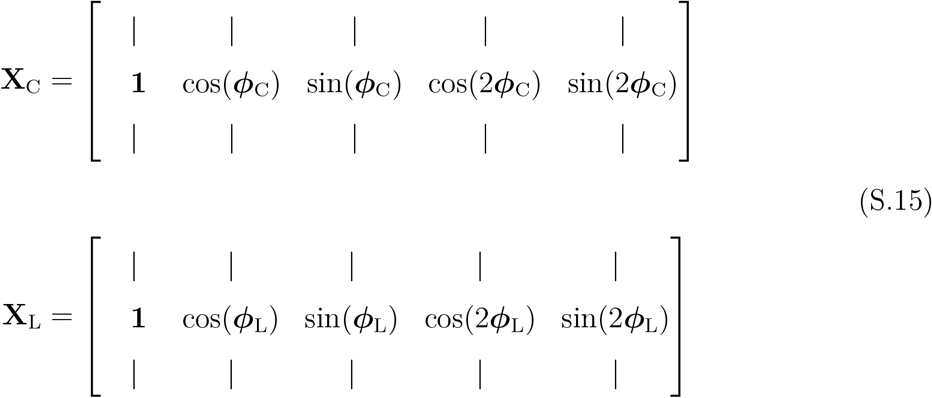

To fit **Y**_C_ and **Y**_L_ to separate 2nd-order Fourier models, the data are modeled as:

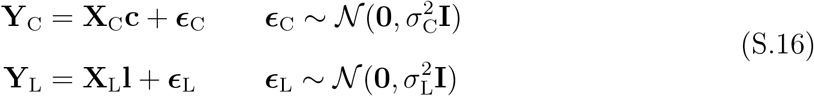

where **c** and **l** are the 2nd-order Fourier coefficients for controls and labels, respectively. The terms ***ϵ***_C_ and ***ϵ***_L_ represent measurement noise, which we assume are represented by independent and identically distributed (i.i.d.) normal random variables with variances 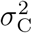 and 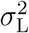, respectively. (Normality of noise in MRI magnitude images is a reasonable approximation when the SNR of a given voxel is sufficiently high. The noise begins to resemble a normal distribution at SNRs as low as ∼3^3^. The control and label data of this study comfortably lie within the high SNR regime, with empirical voxel-wise SNRs on the order of ∼50 for cortical gray matter voxels.) The coefficients **c** and **l** are then estimated via least squares:

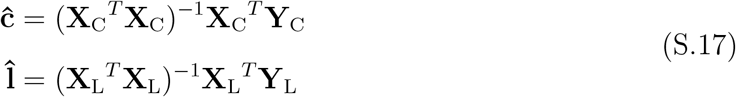

with **ĉ**= [*ĉ*_0_ *ĉ*_cos,1_ *ĉ*_sin,1_ *ĉ*_cos,2_ *ĉ*_sin,2_ ]^*T*^ and 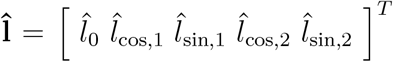 denoting the cor-responding estimates. The difference (signal *S*) coefficients are then computed by control-minus-label subtraction:

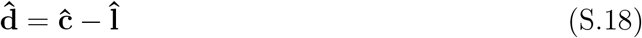

where 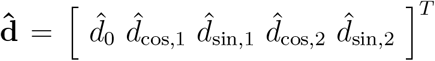. The signal curve *S*(*ϕ*) can then be obtained by inserting the coefficients into a 2nd-order Fourier model (Equation 8 of the main text, reproduced below):

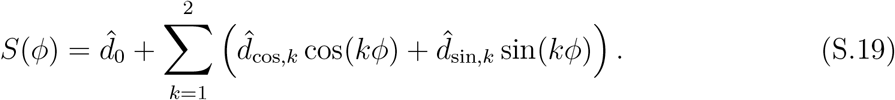

(Equivalently, the control and label coefficients could be inserted into separate 2nd-order Fourier models, followed by subtracting the models.) To compute pulsatility, *S*(*ϕ*) are evaluated over a fine grid of *ϕ* values (for example, *ϕ* ∈ [0, 2*π*] in steps of 2*π* · 0.0001), and then PI can be numerically calculated with Equation 4 of the main text as:

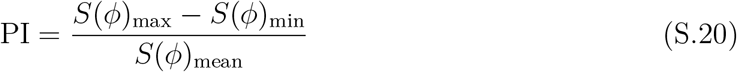

The stability of the PI measurements can be assessed using a residuals permutation approach. We first computed the data estimates 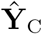 and 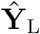 as:

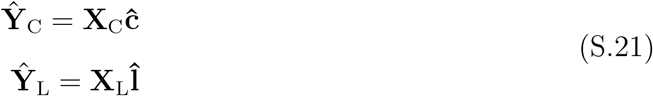

and then computed the residuals 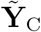 ^C^ and 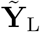 as:

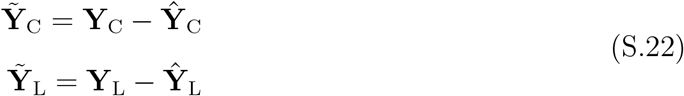

Then, we randomly permuted the residuals in time (separately for labels and controls) and added them back to the fitted data to create simulated data vectors **Y**_C,*Perm,i*_ and **Y**_L,*Perm,i*_:

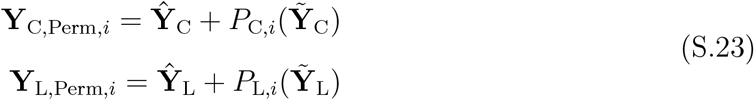

where *P*_C,*i*_(*·*) and *P*_L,*i*_(*·*) denote the permuting/shuffling operators for the control and label data, respectively, for the *i*th iteration. Using these simulated data vectors, PI was computed again following Equations S.17-S.20. This procedure was repeated over 1000 iterations to generate a distribution of PI values, which were then used to compute 95% confidence intervals.

### S3 Impact of Measurement Noise on Pulsatility

This section assesses the bias and SNR of the PI measurement under measurement noise (i.e. noisy control and label data points). We derive the theoretical relationship between measurement noise and the distributions of the estimated difference coefficients 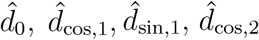 and 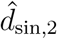 (which were estimated in Equation S.18). The impact of noise on 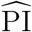 is then assessed via simulations and theoretical approximations of the SNR of PI.

#### S3.1 Theory

##### S3.1.1 SNR and Bias of Pulsatility Index

We first approach this problem by assuming ground truth 2nd-order Fourier coefficients for the control, label and difference signals as follows:

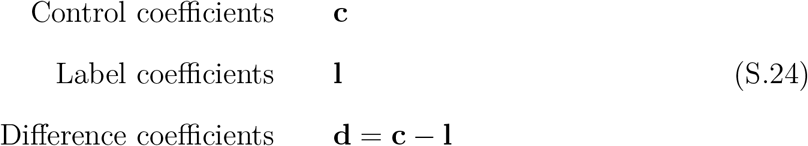

respectively. Following the framework outlined in Section S2, we assume a VSASL scanning experiment that has acquired (*N ×* 1) control and label data vectors **Y**_C_ and **Y**_L_ which follow the model described in Equation S.16. By substituting the expressions of **Y**_C_ and **Y**_L_ (Equation S.16) into the least squares coefficient estimates **ĉ** and 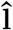 (Equation S.17), we obtain:

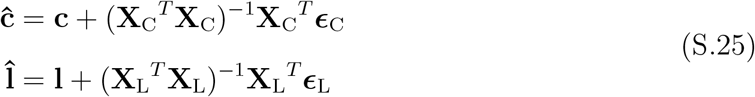

These expressions can then be substituted into Equation S.18 to obtain our estimate of difference coefficients 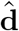 as:

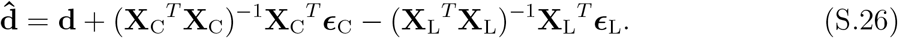

In a realistic experiment, the entries of ***ϕ***_C_ (and ***ϕ***_L_) have values in [0, 2*∈*] that are unevenly spaced across the interval. However, when we have a sufficient number of measurements (e.g. *N* = 72 as in our experiments), approximating ***ϕ***_C_ (and ***ϕ***_L_) as vectors with uniformly spaced entries has negligible impact on the **ĉ** (and 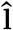) estimation (results not shown). This evenly-spaced approximation is:

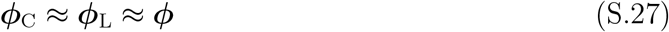

whose entries are given by:

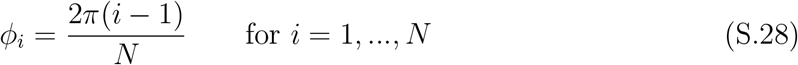

Consequently, the design matrices are also approximated as:

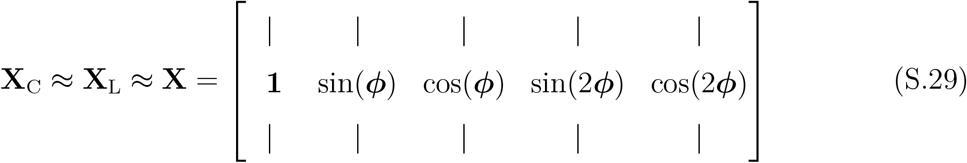

and Equation S.26 simplifies to:

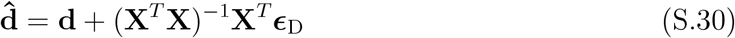

where 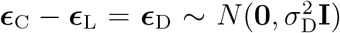 and 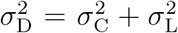. By assessing the expected value and covariance, we see that 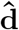 can be described as a normally distributed random vector:

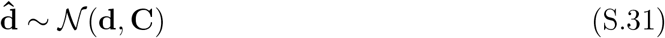

where

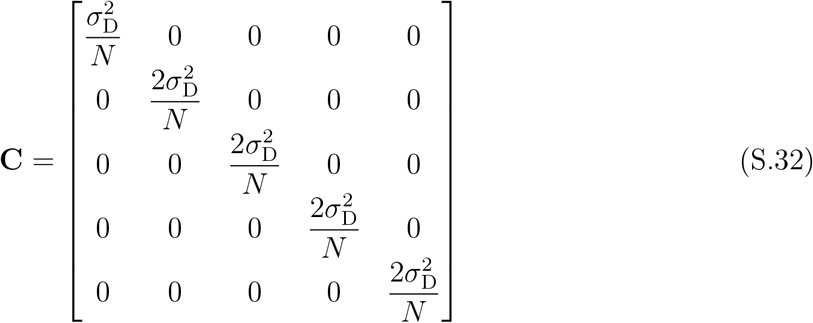

with off-diagonal terms equal to 0 due to the orthogonality of the evenly-spaced 2nd-order Fourier regressors.

This expression of 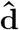 can then be used to examine the behavior of the pulsatility index measurement. Inserting the coefficients into *S*(*ϕ*) yields the estimated VSASL signal curve (Equation 8 of the main text, reproduced below):

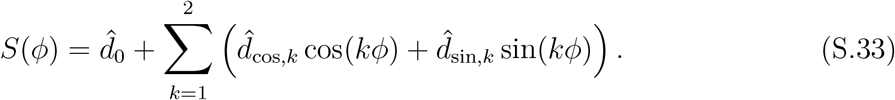

Using similar arguments as in the derivation of Equation 5 of the main text to neglect second-order terms, the pulsatility index estimate 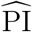 is then calculated via Equation 4 of the main text as:

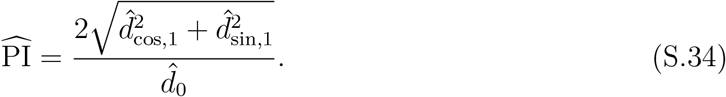

The SNR of 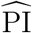 is then defined as:

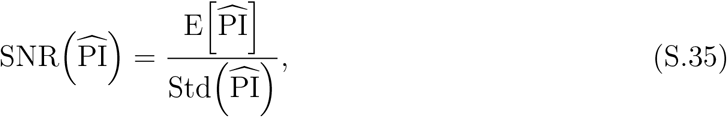

where 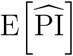 is the expected value and 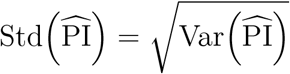. The bias of 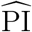 is defined as:

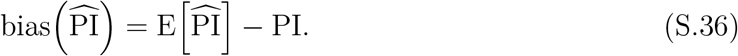

##### S3.1.2 Approximation of SNR

When examining 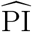 (Equation S.34), its numerator can be identified as a Rician random variable which we denote as 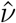:

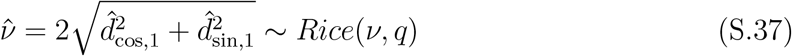

where 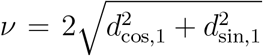 and 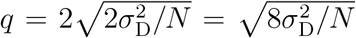 denote the non-centrality and scale parameters of the distribution, respectively. On the other hand, its denominator can be identified as a normal random variable as described by Equation S.32

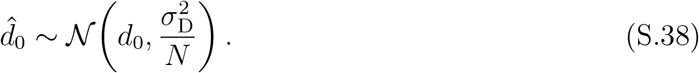

Thus, the pulsatility estimate of

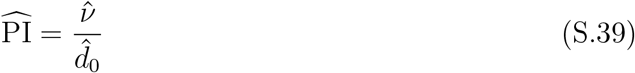

is the ratio of a Rician random variable and a normal random variable. To our knowledge, this exact distribution has not been analytically described in prior literature. However, through several simplifications, we will derive an approximation of 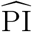 that facilitates the PI SNR model presented in Equation 6 of the main text. The subsequent equations will focus on the derivations, and the errors associated with each step will be examined further in the Methods and Results.

To illustrate these approximations, we start with a random variable *Z* defined as the ratio of *X* and *Y*

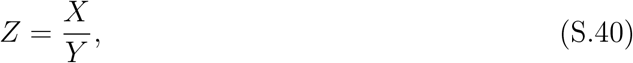

where *X* ∼ *Rice*(*µ*_*x*_, *σ*_*x*_) has a non-centrality parameter *μ*_*x*_ and scale parameter *σ*_*x*_, and 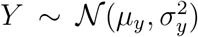 has a mean *µ*_*y*_ with standard deviation *σ*_*y*_. Provided a sufficiently high value of *µ*_*x*_*/σ*_*x*_ (e.g. *>* 5), the Rician random variable *X* can be approximated as a normal random variable 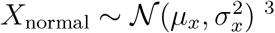, which yields a ratio of two normal random variables. Provided a sufficiently high value of *µ*_*y*_*/σ*_*y*_ (e.g. *>* 10 ^4,5^), which makes the denominator *Y* unlikely to observe negative values, the ratio of two independent normal random variables can be approximated as a single normal random variable^5^ of the form:

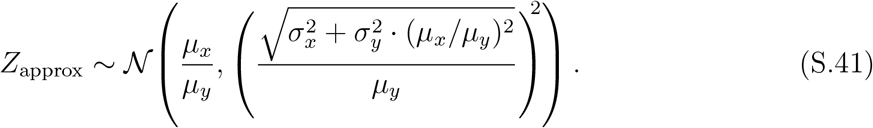

This approximation can be applied to 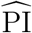 by identifying the corresponding terms 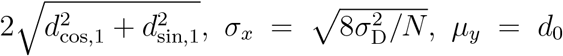 and 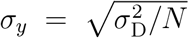. Before applying the 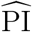 approximation, we first re-express these corresponding terms to elucidate their dependence on *τ*. To do so, we compare the Fourier model of *S*(*t*):

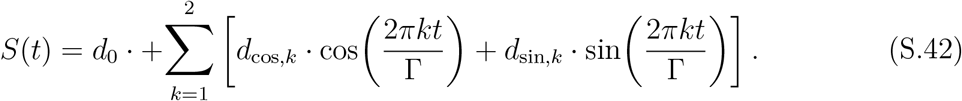

with the expanded form of Equation 3 of the main text:

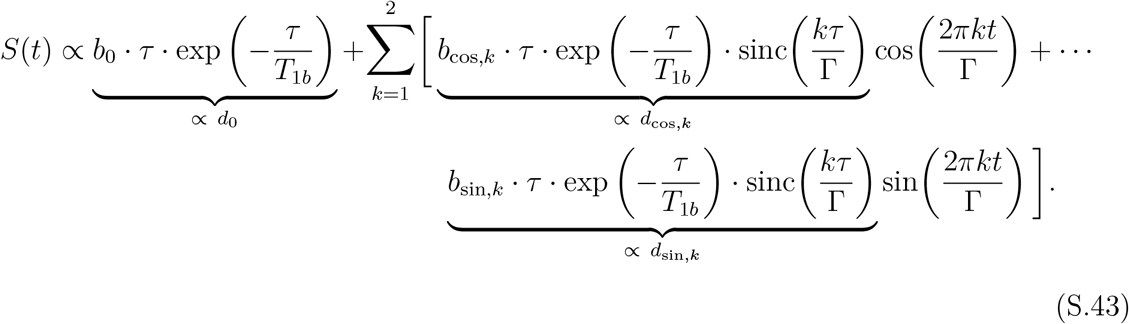

(Note that Equation S.42 is consistent with Equation 8 of the main text by using the substitution *ϕ* = 2*πt/*Γ.) As indicated above by the underbraces, this comparison yields the following definitions for *d*_0_, *d*_cos,*k*_ and *d*_sin,*k*_:

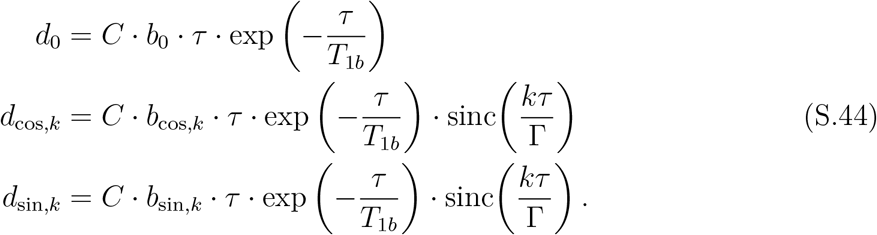

where *C* is the constant of proportionality. Using these definitions for *d*_0_, *d*_cos,1_ and *d*_sin,1_ (with the latter two obtained by setting *k* = 1), the terms *µ*_*x*_, *σ*_*x*_, *µ*_*y*_ and *σ*_*y*_ can be reexpressed as:

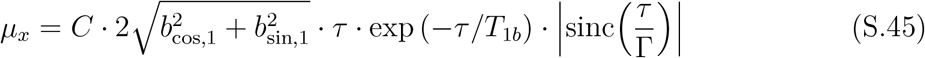

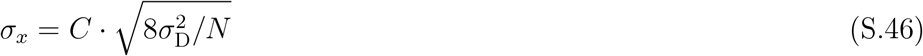

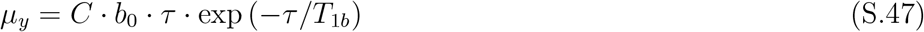

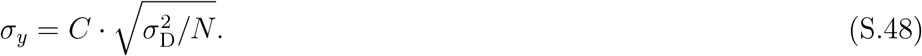

Finally, these terms are applied to Equation S.41 to yield the 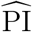 approximation:

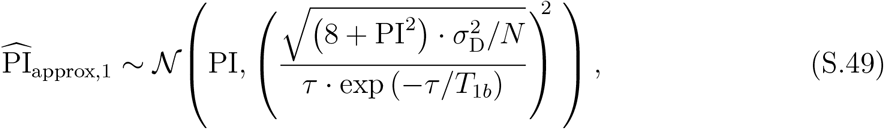

where 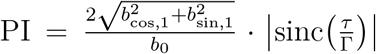 per Equation 5 of the main text (and the subscript ”approx,1” denotes the first of two approximations, with the second coming below). Using Equations S.45-S.48, Figure S3 serves to qualitatively show the regimes of *τ* where the approximations used in Equation S.41 (based on the ratios *µ*_*x*_*/σ*_*x*_ and *µ*_*y*_*/σ*_*y*_) generally hold.

Computing the SNR of 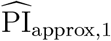 (via Equation S.35) yields:

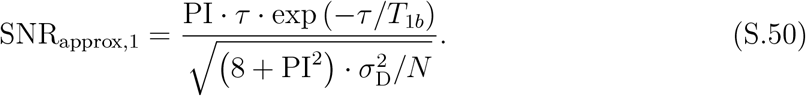

By examining the 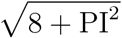 factor (which is present in the standard deviation of 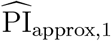 and the denominator of SNR_approx,1_) in a plausible physiological regime, we can make a further simplification. Assuming values of *b*_0_ = 1 and 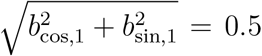 (as in SectionS1.2), the 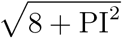 factor simplifies into 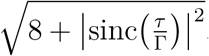. Note that the 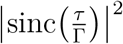 term has a maximum value of 1 (at *τ* = 0 s) that quickly approaches 0 with increasing *τ* due to its envelope of (Γ*/ πτ*)^2^, thus representing a negligible contribution to the overall value of the factor. For example, assuming Γ = 1 s, neglecting 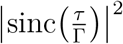 at values of *τ* = 0*/*0.5*/*1.5 s produces errors of 0.057*/*0.038*/*0.013 when expressed as a fraction of 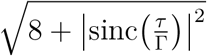 respectively. As empirical support, one can also note that the measured values of PI were all on the order of 1 or less (Figs. 3 and 7). Thus, for a reasonable physiological regime, we can further approximate 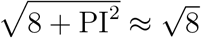, which yields another approximation of 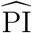:

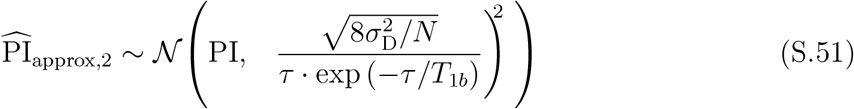

and analytical model of the SNR:

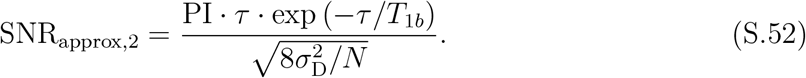

Note that by expanding out the 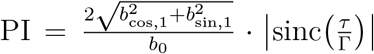 term, this final expression yields the SNR model presented in Equation 6 of the main text (reproduced below):

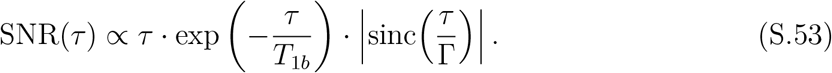

Thus, Equation S.52 is also optimized at *τ*_opt_, given by Equation 7 of the main text.

#### S3.2 Methods

Using the theory derived in Section S3.1.1, we use simulations to assess the impact of noise on the bias and SNR of PI measurements, particularly as a function of *τ*. To do so, we (1) assume a noiseless reference PI(*τ*) curve, (2) add measurement noise, (3) compute distributions of 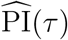 at every *τ* value, and (4) assess the bias and SNR of PI(*τ*) as a function of *τ*.

We begin by assuming reference CBF(*t*) coefficients *b*_0_, *b*_cos,1_, *b*_sin,1_, *b*_cos,2_ and *b*_sin,2_. We assume similar values as in Section S1 (the sinc model error simulations) by using *b*_0_ = 1 and 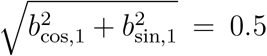, with *b*_cos,2_ = *b*_sin,2_ = 0 also assumed to neglect the 2nd-order component for simplicity. The 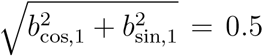 constraint was satisfied by specifying 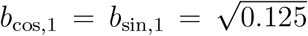 (any pair of values satisfying the constraint will yield equivalent results).

Then, over a grid of *τ œ* [0.001, 2.000] s in steps of 0.001 s, the corresponding *S*(*t*) coefficients **d** = [ *d*_0_ *d*_cos,1_ *d*_sin,1_ *d*_cos,2_ *d*_sin,2_ ]^*T*^ are computed for every value of *τ* based on Equation S.44 using a cardiac period Γ = 1.0 s, and *T*_1*b*_ = 1.6 s (with the proportionality constant *C* arbitrarily set to 1 for simplicity, since *C* cancels out in the eventual PI calculation). Once computed, *d*_0_, *d*_cos,1_, *d*_sin,1_, *d*_cos,2_ and *d*_sin,2_ are inserted into the *S*(*ϕ*) 2nd-order Fourier model (Equation S.19) and then PI is computed (Equation S.20). For example, at *τ* = 500 ms, the coefficients are computed as *d*_0_ = 0.366, *d*_cos,1_ = *d*_sin,1_ = 0.0825 and *d*_cos,2_ = *d*_sin,2_ = 0, which produce a pulsatility value of PI = 0.636. By repeating this procedure for every *τ* value, the ground truth (reference) PI(*τ*) curve is obtained.

Next, we add noise to the simulations and compute 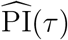. In order to add a realistic amount of noise, we used the in vivo GM ROI *τ* = 500 ms scan analysis results to choose an appropriate value of 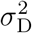. First, 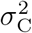 and 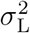 were estimated as the variance of the in vivo control and label fit residuals (Equation S.22) at *τ* = 500 ms, and then 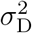 was computed as 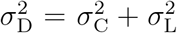. Then, the ratio of *σ* _D_*/d*_0_ was estimated to be *≈* 0.10 (on average across subjects). Since *d*_0_ = 0.366 at *τ* = 500 ms in these noise simulations, we chose *σ* _D_ = 0.0366 to produce an equivalent ratio of *σ* _D_*/d*_0_. This value of *‡*_D_ = 0.0366 (i.e. noise level) was then kept constant across all values of *τ*, consistent with the empirical observation that the residuals at *τ* = 750*/*1000*/*1250*/*1500 ms were comparable in magnitude to the residuals at *τ* = 500 ms.

The number of measurements was set at *N* = 72. The covariance matrix **C** was computed using Equation S.32, and then 10^5^ realizations of 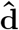 were simulated following 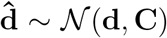 (Equation S.31). We then computed 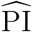 for each realization (Equation S.20) to produce a distribution of 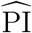 values at every value of *τ*.

We then evaluated 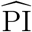 based on its SNR (Equation S.35) and bias (Equation S.36) at every value of *τ*, with the SNR curve then numerically assessed for an optimal value of *τ* :

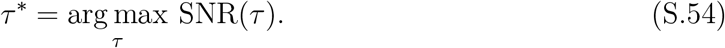

The analytical expressions of SNR_approx,1_ (Equation S.50) and SNR_approx,2_ (Equation S.52) were also evaluated at every value of *τ*, and similarly assessed for their optimal values as well:

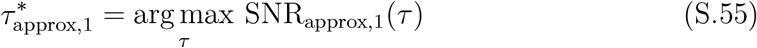

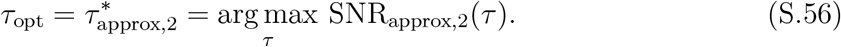

#### S3.3 Results

In Figure S4A, we see generally good agreement between the noiseless reference curve PI and the expected value 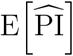, with little spread in 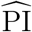 as indicated by the shaded areas. However, 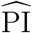 becomes less accurate as *τ* approaches 0 s and the noise begins to dominate. While both the numerator and denominator of the calculation are affected by noise, a noisy denominator 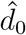 can be particularly problematic for the 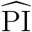 calculation, as individual realizations of *d*_0_ can approach 0 (or even cross into negative values) and make the 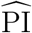 quotient approach infinity. As a result, 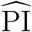 can become unstable in very low-*τ* regimes, which is not surprising due to the low mean signal.

In Figure S4B, the asymptotic behavior of the bias 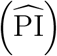 and Std 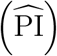 curves for *τ ⟶* 0 reflects the aforementioned stability issues. Another defining feature is the spike in bias around *τ* = Γ (and *τ* = 2 Γ). Here, the reference 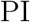 is 0, but 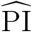 values remain strictly positive due to the *S*_max_ − *S*_min_ numerator of Equation 4, resulting in the positive bias. However, the bias is otherwise negligible for most values of *τ*, including regimes around *τ*_opt_.

Figure S4C shows the simulation-based SNR curve (Equation S.35) alongside SNR_approx,1_ (Equation S.50) and SNR_approx,2_ (Equation S.52), demonstrating excellent agreement in their overall shapes. Furthermore, their respective maxima occur at very similar values of *τ* (represented by the vertical lines). Of note, the difference between SNR(*τ*_opt_) and SNR(*τ* ^*ú*^) is less than 0.1% (18.89 vs 18.90), showing that *τ*_opt_ indeed approximately maximizes SNR. Altogether, the agreement in curve shapes and negligible difference between SNR(*τ*_opt_) and SNR(*τ* ^*\**^) supports the use of the simple theoretical model (Equation 6) to describe the *τ* - dependence of SNR and guide the selection of an optimal *τ* value.

Examining the curves more closely, SNR_approx,1_ is nearly identical to SNR except around *τ* = Γ and *τ* = 2 Γ. The difference in these regimes is expected, as the assumption of normality of the 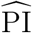 numerator, built into Equation S.41 (and thus into SNR_approx,1_), diverges from its Rician nature in these low-SNR regimes^3^. Figure S5 serves as a supporting figure by comparing the simulated 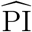 distribution (blue histogram) and the analytical PDF of 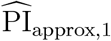 (purple solid line), which approximates the 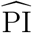 distribution poorly at *τ* = Γ and *τ* = 2 Γ, but otherwise shows excellent agreement for the *τ* values shown. Examining SNR_approx,2_, its curve (green dotted line) is nearly identical to SNR_approx,1_ but is observed to be slightly higher (noticeable in the regime around 0.2 s to 0.6 s) due to the PI^2^ term that was neglected in the derivation step from Equation S.50 to Equation S.52.

### S4 T_1_-weighted Structural Scan Parameters

Two *T*_1_-weighted structural scan configurations were used with similar parameters and both based on a Magnetization-Prepared Rapid Acquisition Gradient Echo (MPRAGE) sequence. The main difference was the acceleration factor, with one using 4*×*-acceleration for shorter scan time, and the other using 2*×*-acceleration to maintain higher SNR for a separate project. The parameters for the former configuration (with the latter configuration noted in parentheses wherever different) were: resolution = 0.9375 mm isotropic (1 mm isotropic), matrix size = 176 *×* 256 *×* 256, 176 slices in the sagittal orientation, flip angle = 8 degrees, TR = 2300 ms (2500 ms), TE = 2.29 ms (2.88 ms), inversion time = 900 ms (1060 ms), GRAPPA = 4*×* (2*×*) acceleration in the A-P phase-encode direction, acquisition time = 3:08 (7:30).

### S5 Iterative Denoising

This section provides supporting details for the iterative denoising method referenced in the main text Section 4.3, which was adapted from Power et al.^6^ with a few modifications. Overall, the algorithm involves identifying outlier volumes over successive iterations, followed by a final censoring of outlier timepoints and subtraction of nuisance variance from the data.

An initial temporal censoring mask was constructed by identifying volumes with framewise displacement (FD) exceeding 0.75mm (with FD computed as in^6^), and volumes with a cardiac period Γ more than 3 scaled median absolute deviations away from the median of the Γ timecourse. The outliers in Γ often corresponded to index finger motion, which distorted the PPG trace and caused unreliable retrospective gating.

For each iteration, the algorithm (1) censors timepoints identified by the (current) temporal censoring mask from the original data and regressors, (2) regresses out nuisance variance and (3) updates the temporal censoring mask with additional volumes identified based on the (current) regression residuals. In our implementation, nuisance regressors were constructed (motion timecourses, motion first-derivative timecourses, and first- and second-order low-frequency drift). To preserve cardiac-driven fluctuations, we also included 2nd-order Fourier regressors of controls and labels, which were constructed by starting with **X**_C_ and **X**_L_ (derived in Equation S.15 in SI Section S2) and interleaving with 0’s to account for the alternating control/label ASL acquisition scheme. For example, 0’s were inserted into the **X**_C_ regressors for every row corresponding to a label data point (and vice versa for the **X**_L_ regressors at control data points). Figure S6A shows how they are interleaved with 0’s and then concatenated alongside the nuisance regressors. Figure S6B shows **X**_C_ and **X**_L_ separately and sorted in cardiac phase *ϕ* order to better illustrate the shape of these regressors.

The nuisance and Fourier regressors were both input into the iterative denoising algorithm, and additional timepoints were censored with each iteration based on DVARS and SD metrics as in the original Power et al. paper^6^. The algorithm was stopped when no new timepoints were censored for a given iteration. Then the nuisance regressors were orthogonalized with respect to the Fourier regressors, and the orthogonalized nuisance fits were subtracted out.

## S7 Figures

**Figure S1:**
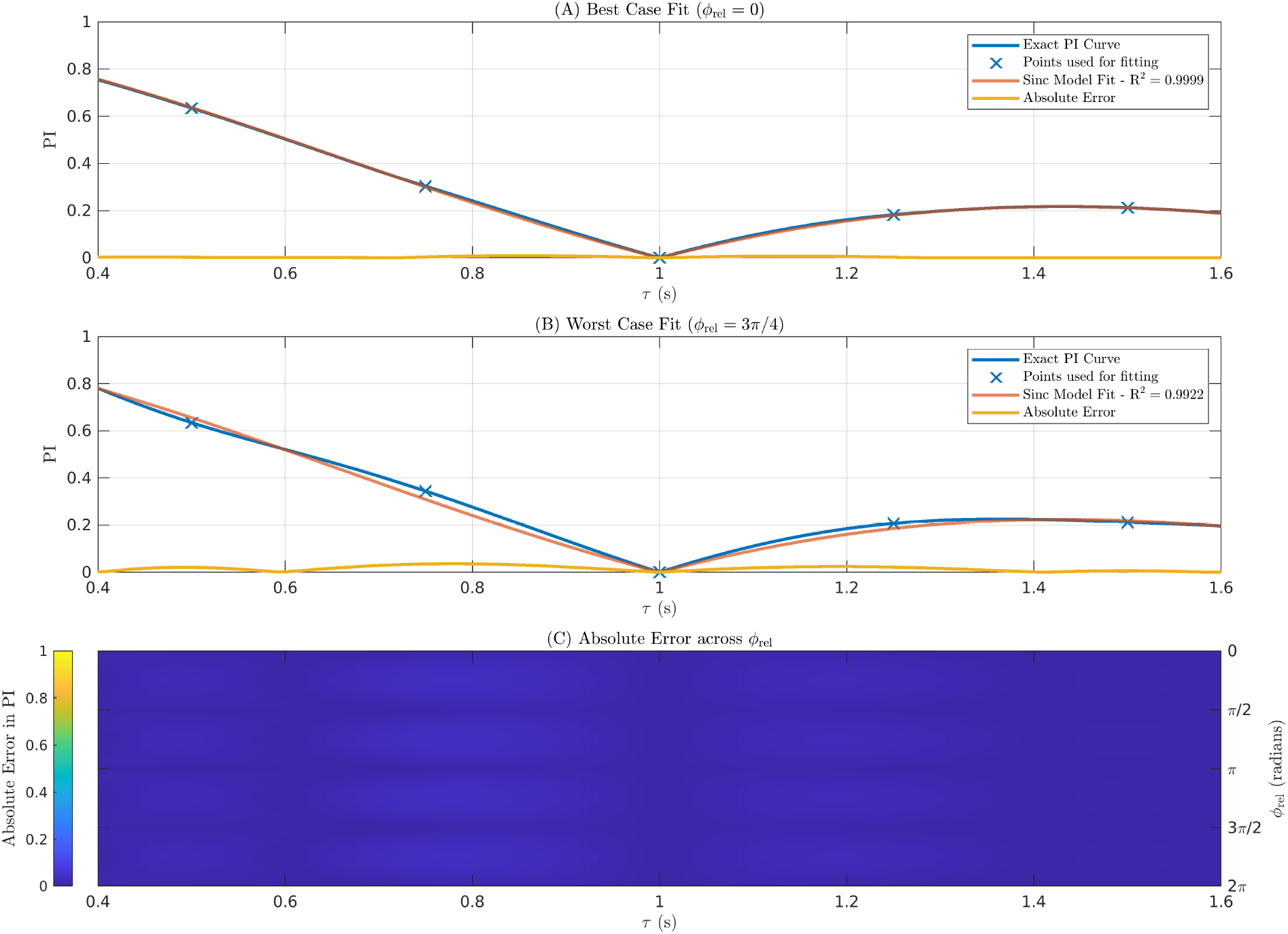
Evaluating Sinc Approximation Error: (A) Best case fit of Equation 5 to the exact PI(*τ*) curve. The curves are nearly identical, with negligible absolute error (*<* 0.01) over all values of *τ* ∈ [0.4, 1.6] s and R^2^ = 0.9999. (B) Worst case fit of Equation 5 to the exact PI(*τ*) curve. Even in this worst case, the agreement between curves is excellent, with minimal absolute error (*<* 0.05) over all values of *τ* ∈ [0.4, 1.6] s and R^2^ = 0.9922. (C) Image of the absolute error across all values of *ϕ* _rel_ ∈ [0, 2 *π*].

**Figure S2:**
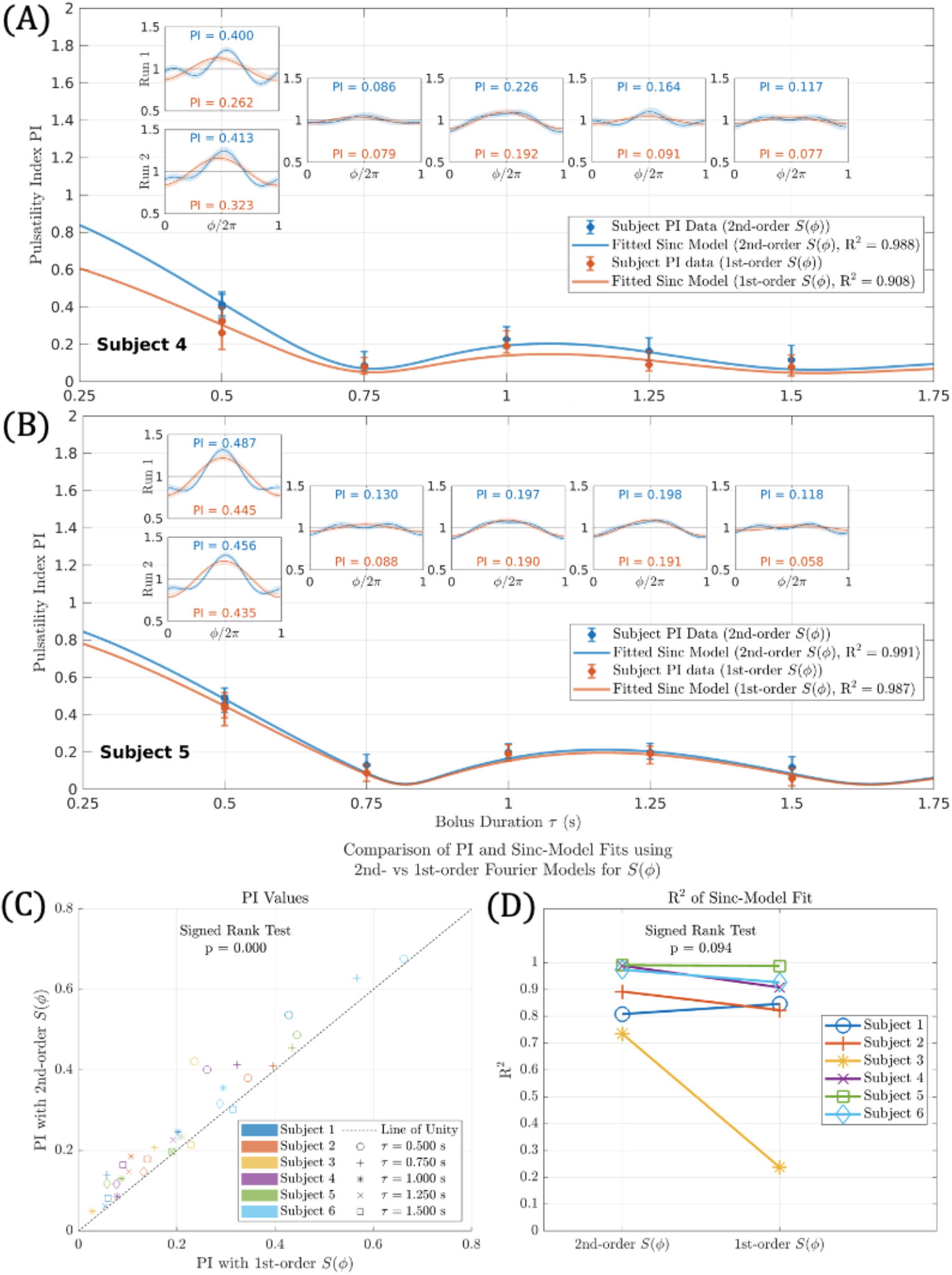
Comparison of the *τ* -stepping results when using 2nd-vs 1st-order Fourier models to describe *S*(*ϕ*). Shown in (A) and (B) are results from two representative subjects (4 and 5). Plotted in blue are the *τ* -stepping results using a 2nd-order Fourier model (identical to those shown in Figure 3 of the main text). Plotted in orange are the PI values when using a 1st-order Fourier model. The insets show comparisons of the *S*(*ϕ*) curves for each approach. Shown in (C) is a scatter plot of PI values, with the 1st-order approach on the x-axis and the 2nd-order approach on the y-axis. Shown in (D) is a comparison of the R^2^ of the sinc model fits.

**Figure S3:**
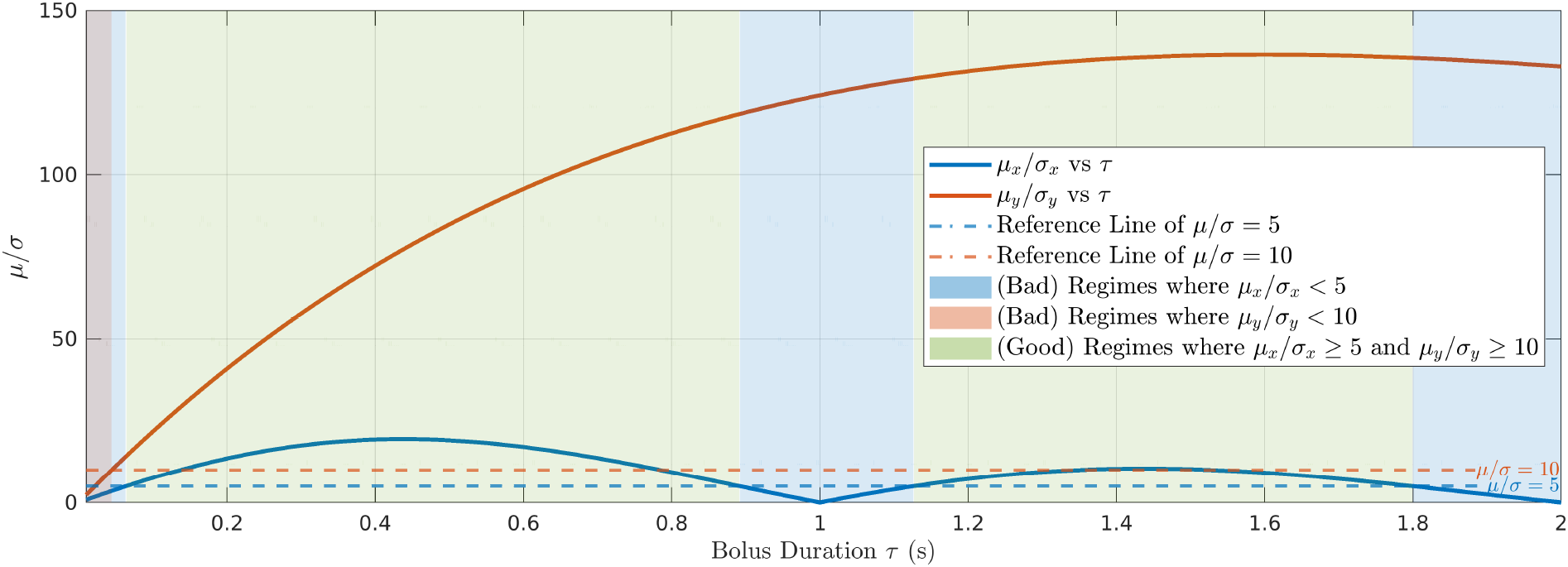
A supporting figure for qualitatively understanding the regimes of *τ* where the approximations used in Equation S.41 generally hold. The expressions of *µ*_*x*_, *σ* _*x*_, *µ*_*y*_ and *σ* _*y*_ were evaluated using Equations S.45-S.48, with values of 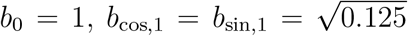, Γ = 1 s, *σ* _D_ = 0.0366 and *N* = 72 that were used in Section S3.2. The blue regions indicate where *µ*_*x*_*/ σ* _*x*_ is less than a threshold of 5, where the normal approximation of the Rician numerator of 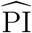 becomes relatively poor. The orange regions indicate where *µ*_*y*_*/ σ* _*y*_ is less than a threshold of 10^4,5^, where the approximation of the ratio of two normal random variables as a single normal random variable becomes relatively poor. The green regions indicate the regimes where both thresholds are exceeded and the approximations hold relatively well. These green areas are consistent with the regimes in Figure S4C showing excellent agreement between SNR and SNR_approx,1_.

**Figure S4:**
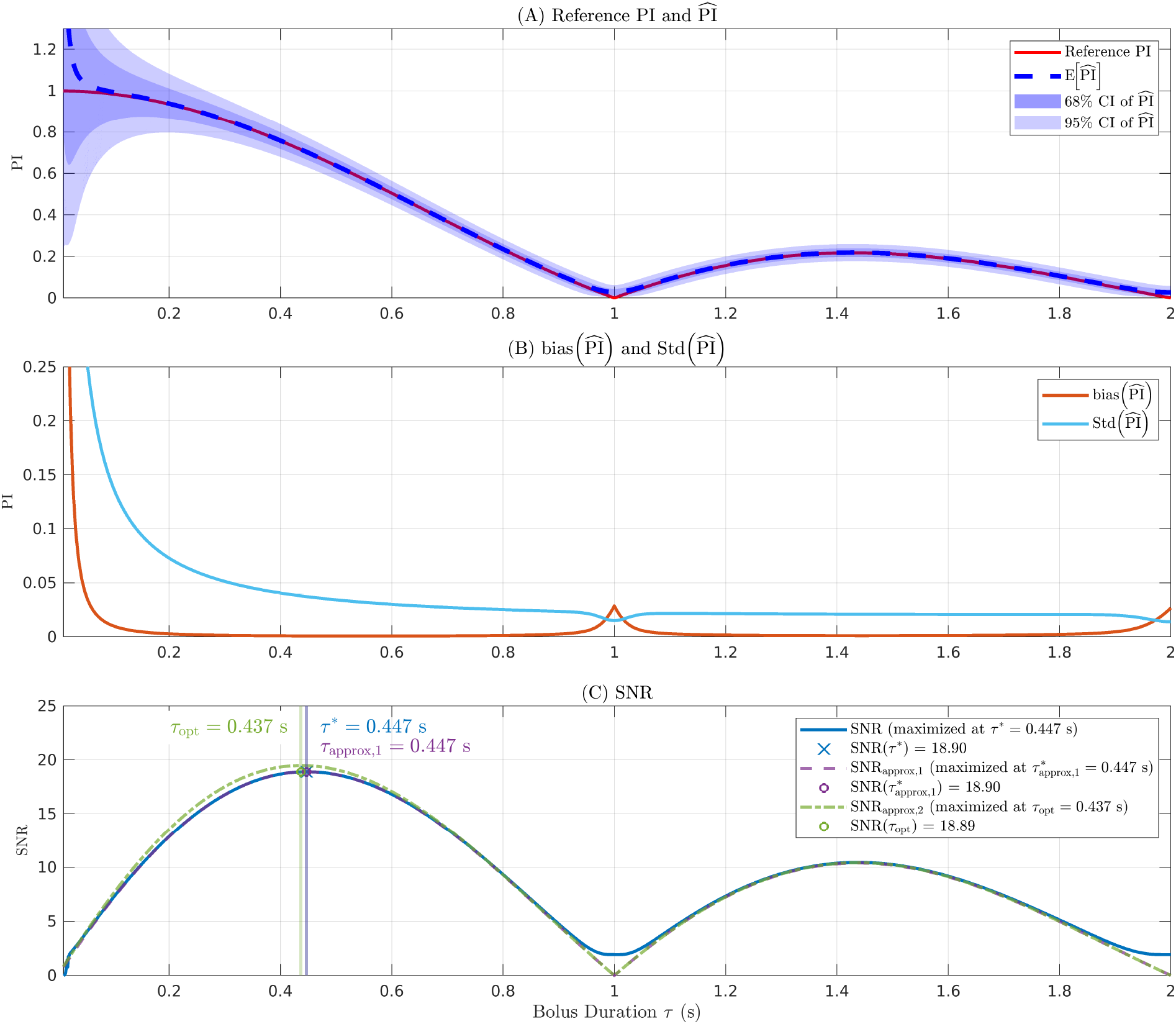
Noise simulations for 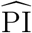. (A) Reference PI curve (red), expected value of 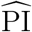 (blue dashed line), and the spread of 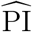 (shaded regions corresponding to 68% and 95% confidence intervals). (B) The bias (orange) and standard deviation (cyan) of 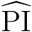 as a function of *τ*. (C) The SNR of the 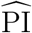 measurement (blue solid line) and approximations SNR_approx,1_ (Equation S.50, purple dashed line) and SNR_approx,2_ (Equation S.52, green dash-dotted line), along with the *τ* values where their maxima occur (i.e. 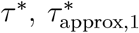 and *τ*_opt_, respectively) denoted by the vertical lines. The plotted points indicate the SNR (blue solid line) evaluated at those *τ* values.

**Figure S5:**
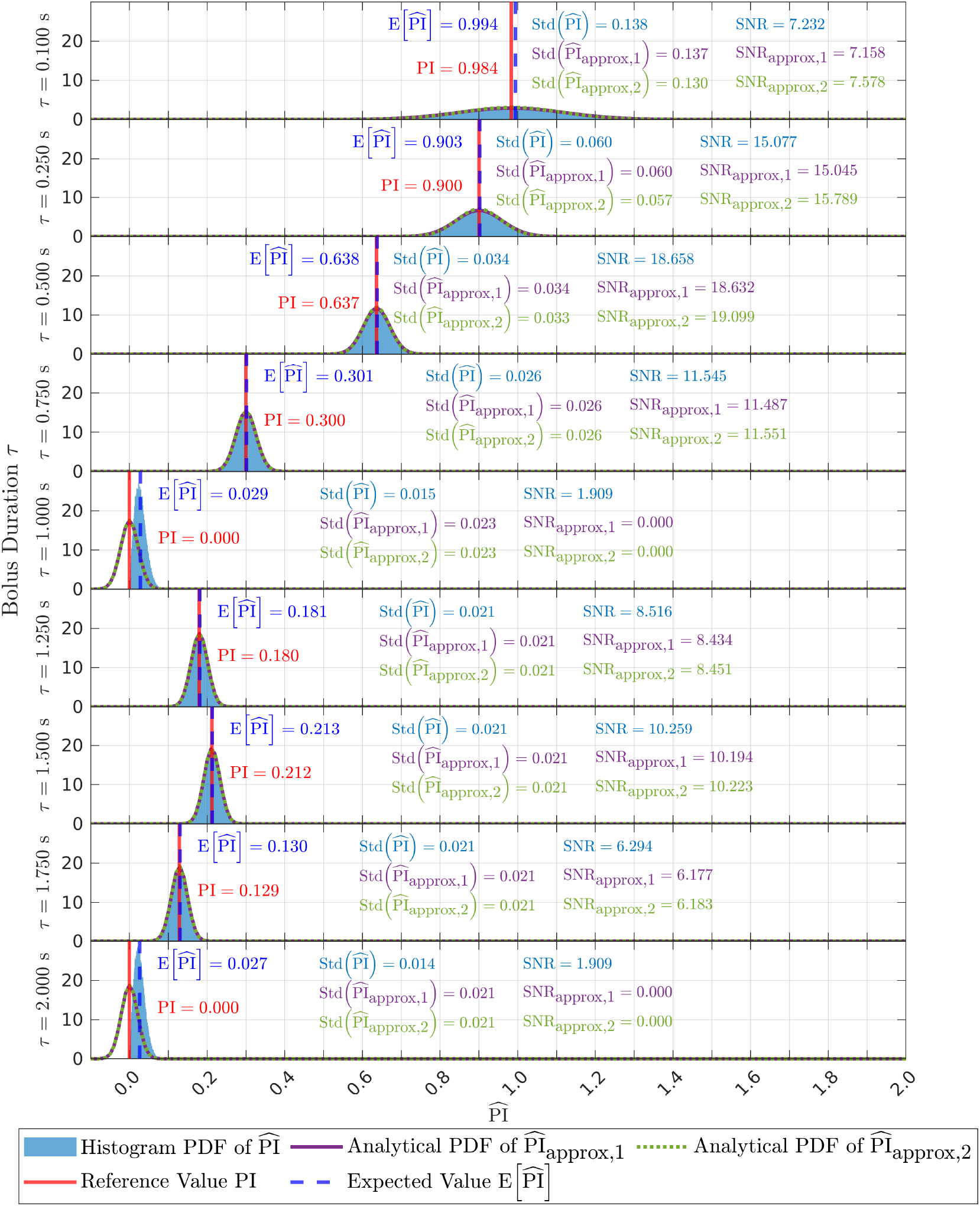
This is a supporting figure for understanding the SNR curves shown in Figure S4C. Shown are the simulated distributions of 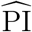 (blue histogram) compared to the analytical PDFs of 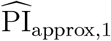 (purple solid line) and 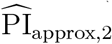 (green dotted line), for a range of selected *τ* values in steps of 0.250 s (except the first step at 0.100 s). Also indicated are the reference PI value (red vertical line) and 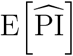 (blue vertical dashed line). At each value of *τ*, the texts show the values involved in computing the SNR. The histogram and its analytical approximations (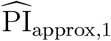 and 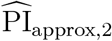) generally agree well, except at *τ* = Γ and 2 Γ, where the built-in normal approximation of the Rician numerator of 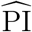 no longer holds.

**Figure S6:**
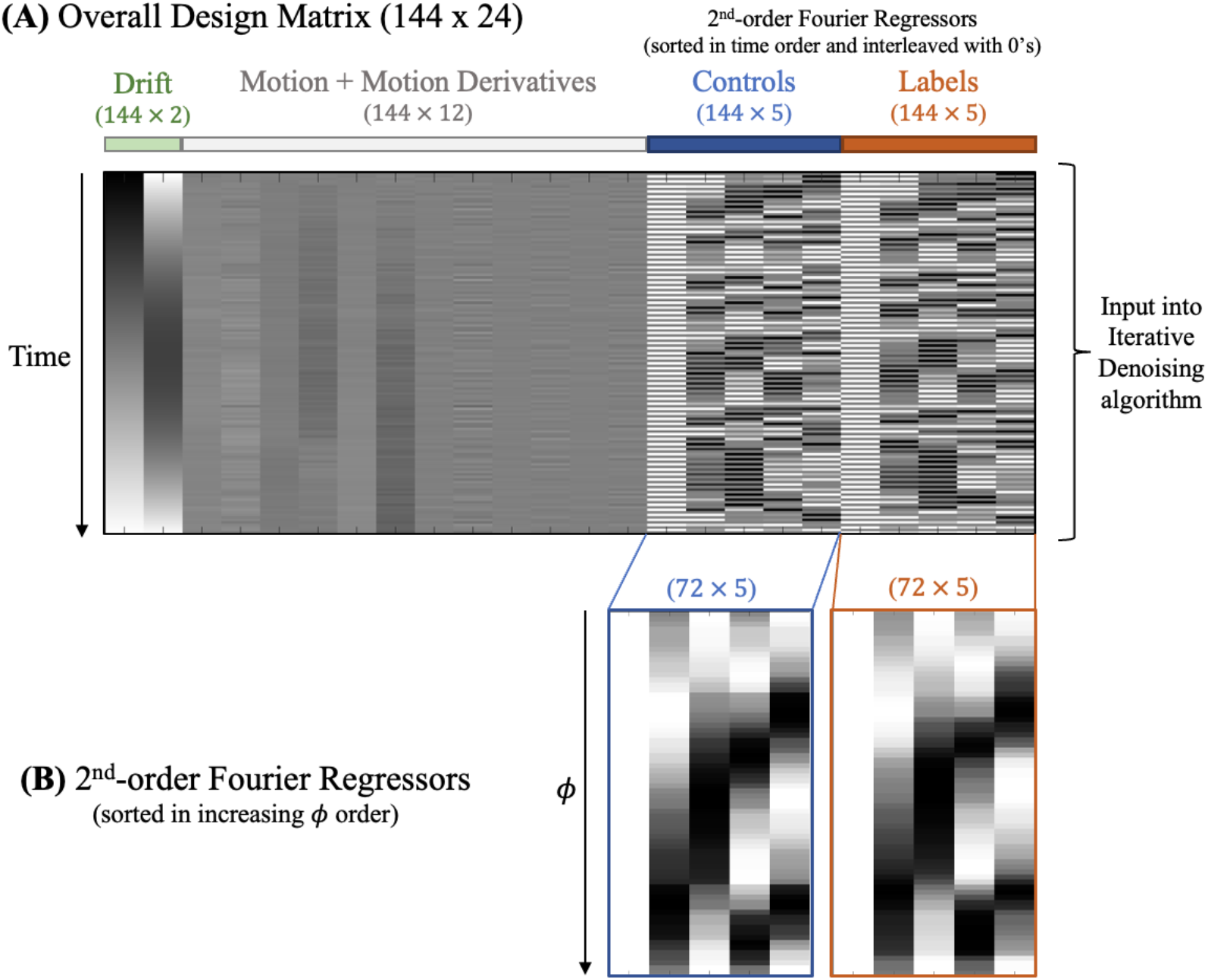
Illustration of the 2nd-order Fourier models in the iterative denoising algorithm. (A) The overall design matrix used in the iterative denoising algorithm of Section 4.3 of the main text. The 2nd-order Fourier matrices, representing separate models for controls and labels are sorted in time according to the temporal acquisition order of the control and label volumes and interleaved with 0’s according to the label/control alternation. They are then concatenated alongside the nuisance regressors (drift, motion and motion derivatives) and input into the iterative denoising algorithm. (B) The 2nd-order Fourier design matrices (constructed as in Equation S.15) separated out and sorted in cardiac phase *ϕ* order for visualization.

